# Inequalities in Healthcare Use during the COVID-19 Pandemic

**DOI:** 10.1101/2023.04.26.23289095

**Authors:** Arun Frey, Andrea M. Tilstra, Mark D. Verhagen

## Abstract

The COVID-19 pandemic has led to severe reductions in non-COVID related healthcare use, but little is known whether this burden is shared equally across the population. This study investigates whether the reduction in administered care disproportionately affected certain sociodemographic strata, in particular marginalised groups. Using detailed medical claims data from the Dutch universal health care system and rich registry data that cover all residents in The Netherlands, we predict expected healthcare use based on pre-pandemic trends (2017– Feb 2020) and compare these expectations with observed healthcare use in 2020. Our findings reveal a substantial 10% decline in the number of weekly treated patients in 2020 relative to prior years. Furthermore, declines in healthcare use are unequally distributed and are more pronounced for individuals below the poverty line, females, the elderly, and foreign-born individuals, with cumulative relative risk ratios ranging from 1.09 to 1.22 higher than individuals above the poverty line, males, young, and native-born. These inequalities stem predominantly from declines in middle and low urgency procedures, and indicate that the pandemic has not only had an unequal toll in terms of the direct health burden of the pandemic, but has also had a differential impact on the use of non-COVID healthcare.

## Introduction

The COVID-19 pandemic (henceforth: pandemic) has affected healthcare services across the world, placing systems under severe strain, contributing to delayed or even missed care and treatment (Cox et al. 2021, Gardner et al. 2020, Li et al. 2020, Mehrotra et al. 2020, Oliver 2021, Shoukat et al. 2020, Whaley et al. 2020). Although overall declines in healthcare use are well recorded (Liao et al. 2020), it remains unclear how this reduction in administered care affects different strata of the population. We know that the direct effects of the pandemic have had an unequal burden on marginalised populations (Apea et al. 2021, Bassett et al. 2020, Cuéllar et al. 2021, Hawkins et al. 2020, Mena et al. 2021, Sesé et al. 2020, Sharma et al. 2021, Jordan et al. 2020), but little is known of how the collateral effects of the pandemic have affected different demographic groups. We address this gap by analysing detailed information on healthcare expenditures and rich registry data on all individuals registered in The Netherlands between 2017 and 2020, quantifying the number of missed healthcare treatments for different population groups.

Beyond elevated rates of mortality from COVID (Aburto et al 2021; Schöley et al. 2022), and the emerging consequences of long COVID (Choutka et al. 2022; Crook et al. 2021; Smith 2022), the pandemic has also disrupted non-COVID healthcare services. Healthcare systems across the world responded to the imminent threat of the pandemic by changing their institutional practices in order to anticipate increased hospitalisations due to COVID, necessitating a decline in the availability of non-COVID healthcare procedures (Li et al. 2020, Oliver 2021, Shoukat et al. 2020). This decline was especially pronounced for procedures deemed nonessential or elective, as hospitals and public health authorities made triaging decisions over which treatments to prioritise (Cox et al. 2021, Gardner et al. 2020, Mehrotra et al. 2020, Whaley et al. 2020). Although healthcare procedures decreased during the pandemic, it is unclear whether this reduction was evenly distributed among all individuals or if specific population groups, such as marginalised communities, were disproportionately affected. Insights into how the decline of non-COVID healthcare differed between demographic groups remain an important line of inquiry that can inform policy efforts to mitigate the long-term effects of missed or delayed healthcare (Dinmohamed et al. 2020; Mehrotra et al. 2021; Oosterhoff et al. 2023; Padmaja and Behera 2023; Riera et al. 2020).

There are various reasons to expect group differences in the decline of non-COVID healthcare procedures. First, the pandemic likely changed the underlying need for certain medical procedures. The pandemic and subsequent lockdown led to drastic behavioural changes that altered pre-existing patterns of illnesses and injuries. Traumatic injuries following traffic accidents, for example, declined considerably during lockdown, as individuals spent less time commuting or travelling (Yasin et al. 2021). Such injuries are more common for some population groups than others, so changes to health needs may have contributed to differences in healthcare use between population groups. The need for other procedures, such as cancer treatments, should have remained largely constant during the pandemic.

Second, people may have avoided visiting the hospital during the pandemic, resulting in missed care (Hafner 2020; Liao et al. 2020; Splinter et al. 2021; Czeisler et al. 2020). In The Netherlands, there are notable patterns in care avoidance during the pandemic: older individuals, females, unemployed people, and people in poorer health reported the highest rates of healthcare avoidance (Splinter et al. 2021). Since the risk of adverse health effects following COVID contraction differs by demographic group, such fears may have disproportionately dissuaded some population groups from seeking the non-COVID care they needed, contributing to differences in healthcare use.

Third, demographic groups may differ in their ability to successfully navigate a healthcare system under pressure. It is already known that marginalised people frequently experience barriers to accessing the same quality healthcare as their more advantaged peers, a pattern that can manifest through structural barriers (e.g., distance to healthcare facilities), financial barriers (e.g., cost of healthcare), or health beliefs and literacy (Levesque et al. 2013; World Health Organization 2010). This is especially the case with private healthcare systems, such as the United States, but inequalities in access have also been documented in public healthcare systems that, in theory, should provide equal access to all individuals (Gerritson et al. 2009; Schoevers et al. 2010; Uiters et al. 2006). Given strong reductions in the availability of non-COVID healthcare, it is likely that similar inequalities materialised when procedures became more scarce and thus harder to access.

In this paper, we quantify the weekly decline in non-COVID healthcare use during the first year of the pandemic (2020) and compare declines across various demographic characteristics. In doing so, we examine whether the decline in healthcare use disproportionately affected certain sub-populations. To do this, we rely on rich population-level data from The Netherlands that we match to the universe of individual healthcare activities in the period 2017–2020 using full population insurance data from the Dutch universal health insurance system. We predict the expected number of individuals using healthcare based on pre-pandemic trends (2017 to February 2020) and compare these expectations with the observed number of individuals using healthcare between March and December 2020, across various population groups. In doing so, we are able to assess both the magnitude and distribution of the decline in healthcare use during the first year of the pandemic for all individuals in The Netherlands.

The Netherlands presents a unique context for analysing inequalities in healthcare use during the pandemic, as healthcare coverage is universal and publicly available to all residents. Combined with The Netherlands’ i) relatively small land mass, high population density, and dense infrastructure, and ii) efficient healthcare system, there is little reason to expect inequalities in access to healthcare services, in principle. In other words, The Netherlands serves as a best case scenario for a resilient and equitable healthcare system. In line with most countries globally, the Dutch government made concerted efforts to catch-up on postponed healthcare during the summer months of 2020—when the number of COVID cases was relatively mild—but subsequent rises in infections shortly thereafter have led to a considerable backlog (van Giessen et al. 2020). At the time of writing, catching up on missed care is still a central policy issue.

Based on pre-pandemic trends in healthcare procedures, we estimate that in the early peak of the pandemic (mid-March to mid-May 2020) there were weeks with more than 300,000 people not receiving care in the Netherlands, which amounts to close to 45% fewer individuals treated. By the end of 2020, there were nearly 3 million fewer patients treated that year than expected, a nearly 10% reduction from prior years. The declines in healthcare procedures are not offset by the number of individuals entering the Dutch healthcare system for COVID, demonstrating the immense strain of the pandemic on the standard provision of healthcare. We also find that although this decline is most pronounced for non-critical procedures, both less-urgent and highly-urgent procedures declined in 2020: there were 12% fewer patients seen for low urgency procedures, compared to 7% for highly urgent procedures.

Importantly, we find strong differences between sociodemographic groups in the reduction in care, particularly for non-critical procedures. Less urgent procedures were more likely to be missed by migrants, impoverished people, women, and the elderly. Although urgent procedures declined equally across most population groups, older age groups were less likely to have urgent procedures compared to younger individuals, indicating a disproportionate impact of the pandemic on this demographic. We also find striking differences across demographic groups when separately assessing medical procedures: declines in trauma were mostly concentrated among young males between the ages of 18 to 29. Reductions in highly urgent oncological care were almost twice as large for women compared to men and considerably larger for the elderly population.

As the direct effects of the COVID-19 pandemic slowly wane, missed or delayed care continues to pose major challenges to healthcare systems across the world, as delayed entry into care can exacerbate severe health burdens in later life (Wilson and Jungner 1968). As efforts to catch up on delayed care are still under way, it is crucial to understand how declines in non-COVID healthcare were distributed across sociodemographic groups and target interventions accordingly. Our quantification of inequalities thus contributes to a growing body of literature on the unequal ways the pandemic has affected population health (Bambra et al. 2020; Mamelund and Dimka 2021) and offers further insight for policymakers and healthcare providers as they strive to catch up on missed care.

## Results

To estimate declines in healthcare use during the COVID-19 pandemic, we calculate the weekly number of unique patients treated in the Dutch healthcare system for the period between 2017 and 2020. We model this weekly count based on data from January 2017 through February 2020 and generate weekly predictions for the remainder of 2020. We then compare the predicted weekly number of treated individuals to the actually observed number (see Methods).^1^ We estimate that on a weekly level, nearly 3 million fewer patients received care by the end of the first year of the pandemic than expected. Following the onset of the COVID-19 pandemic in March 2020, the number of weekly treated individuals declined rapidly (Figure 1). At the height of the reduction at the end of March, 2020, 300,000 fewer individuals received care than expected, a near 45% decline in healthcare use. Although the number of individuals missing treatment decreased from mid-May onward, there was little to no catching up during the summer months, despite comparatively fewer COVID infections during this time and concerted efforts to catch up on missed care (van Giessen et al 2020).

**Figure 1:**
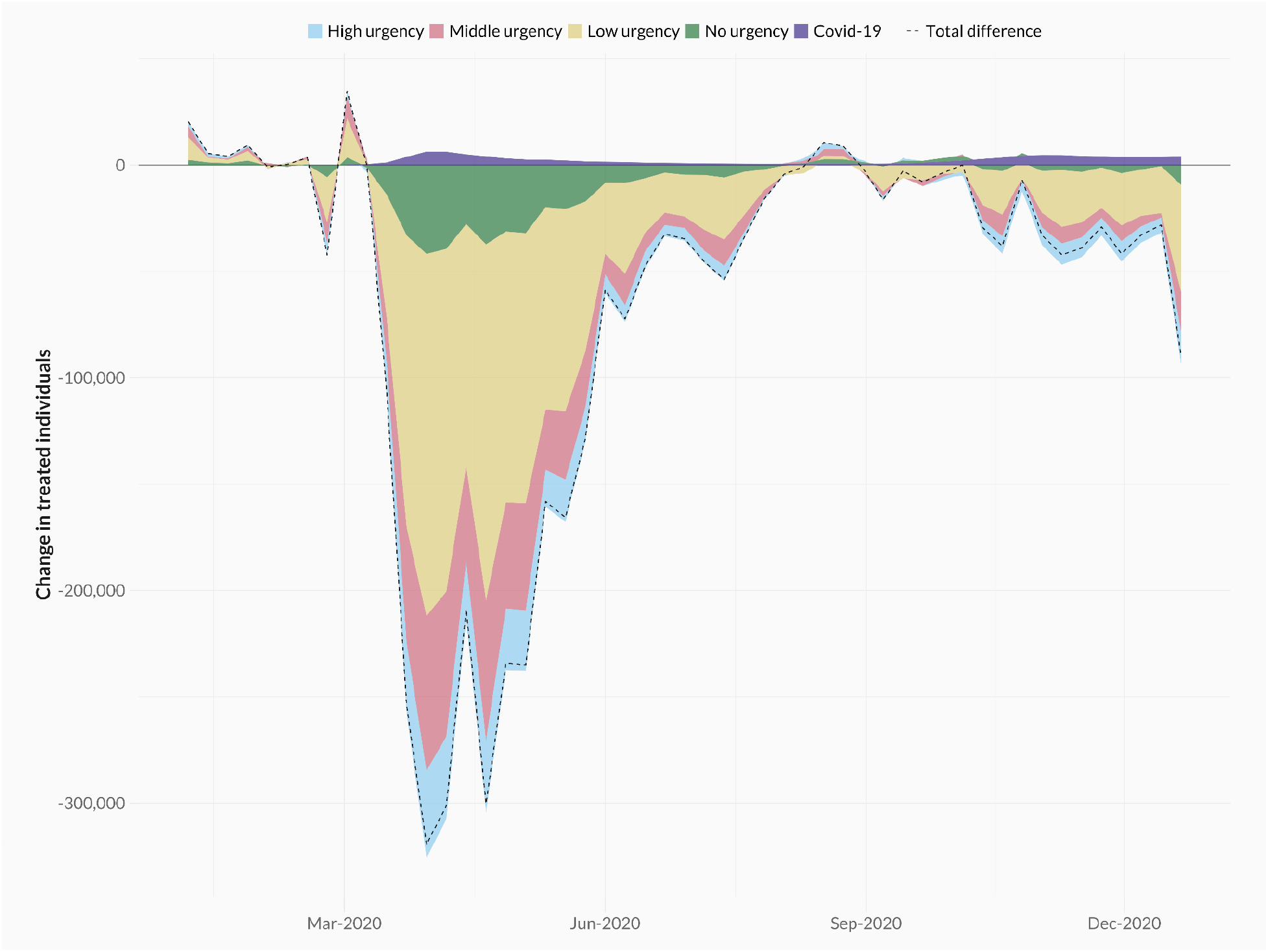
Difference between the observed and predicted number of treated individuals per week in 2020. Colours differentiate between urgency types (high, middle, low, and no urgency) and patients treated for COVID-19.

The observed decline in individuals receiving care following the onset of the pandemic is many times larger than the increase in treatments related to COVID infections. At the height of hospital use from COVID (the week of March 23rd, 2020), there were more than 6,300 additional individuals that received treatment for COVID infections, compared to a simultaneous decline in non-COVID related healthcare of more than 300,000 individuals. We observe a less pronounced decline during the second peak of COVID hospitalisations at the end of 2020, which further underscores the unique challenges of the early weeks of the pandemic.

Most of the decline in healthcare procedures stems from those with low levels of urgency – i.e. procedures that do not have to be performed in the next week. However, we also observe a considerable decrease in highly urgent activities that require medical attention within a week: at the end of 2020, there were 7.6% fewer patients receiving highly urgent care, compared to 12.6% fewer patients receiving low urgency treatments. These declines in year-over-year care are substantial, especially when considering that the pandemic only affected the final ten months of 2020. Much of the observed decline in highly urgent care can be explained as a consequence of strong reductions in trauma-related cases (Figure SI-1).^2^ However, the pandemic also decreased highly urgent oncological care, with 5% fewer patients receiving critical oncological care by the end of 2020 than in previous years, even though the need for oncological procedures should not have changed as a result of the pandemic.^3^

Importantly, the observed decline in non-COVID healthcare use in 2020 was not consistent across sociodemographic groups. In Figure 2, we show the cumulative impact of the pandemic on healthcare use across different demographic groups, which allows us to assess the magnitude of the uneven toll of the pandemic. By the end of 2020, impoverished individuals were 12% more likely to have missed a low urgency procedure and 26% more likely to have missed a medium urgency procedure than their non-poor counterparts (see Figure 3). We document a similar trend for individuals with a migrant background, who were 12% and 10% more likely to miss low and medium urgency procedures than native Dutch individuals. Elderly people and females also saw greater declines in healthcare use than younger people and males. For highly urgent procedures, we find considerably lower levels of inequality, but continue observing a substantial age gradient in urgent health use: individuals aged 76 or above are 20% more likely to miss a highly urgent treatment compared to young patients (18–29).

**Figure 2:**
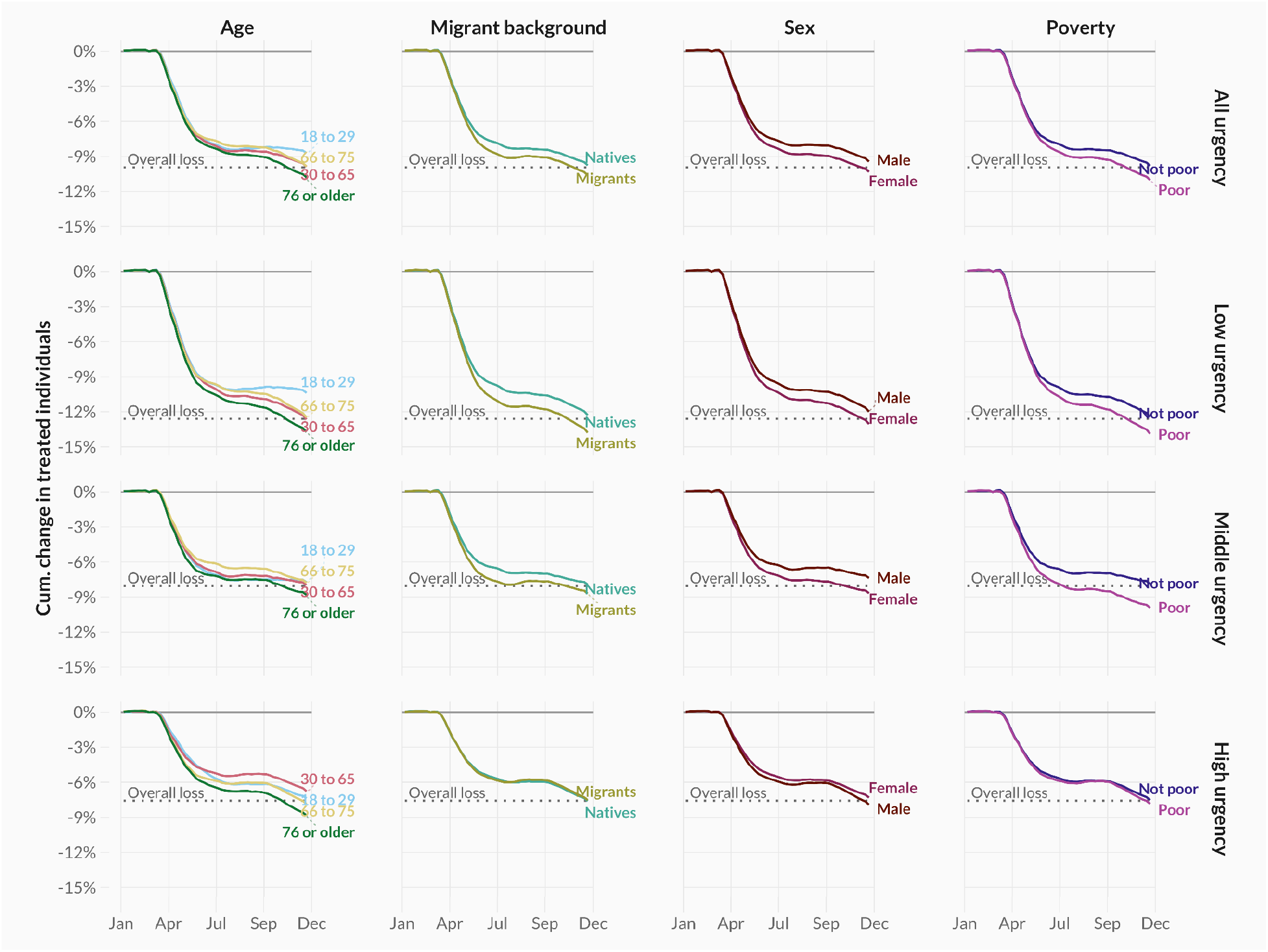
Cumulative age- and sex-adjusted differences between the observed and predicted number of treated individuals in 2020, across urgency types (rows) and demographic groups (columns).

**Figure 3:**
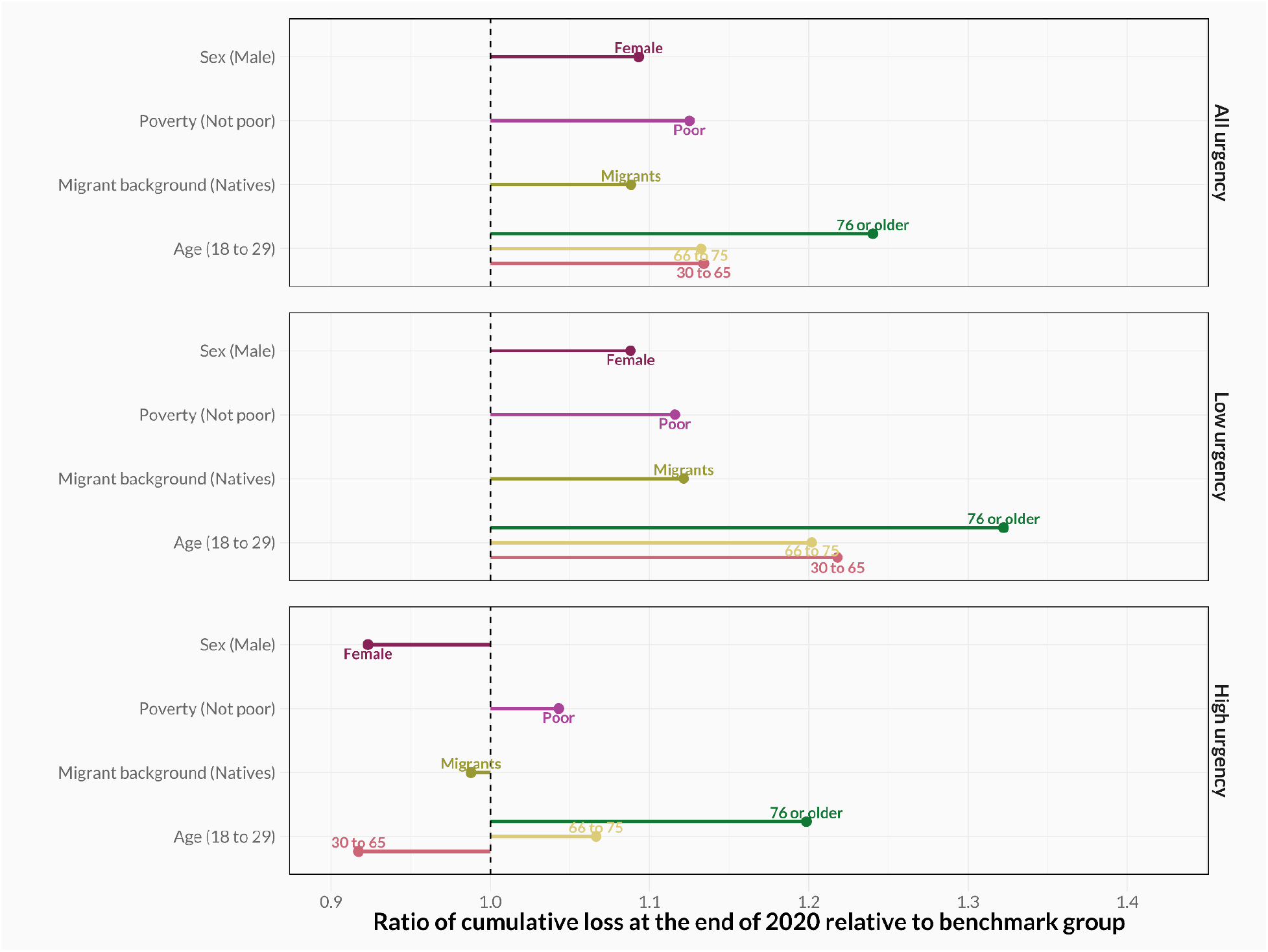
Ratio of the cumulative loss across 2020 for each demographic group relative to a benchmark group (in brackets).

We also explore whether the observed group differences are primarily due to underlying changes in the need for healthcare. To do this, we examine inequalities in the use of oncological (left) and trauma (right) healthcare (Figure 4). While the risk of traumatic injuries reduced considerably during the pandemic, the need for oncological care should, in theory, remain the same. In line with these expectations, we observe stronger overall reductions in trauma-related care than oncological care. In addition, the strongest reduction in trauma-related procedures can be observed among young males, who are otherwise less impacted by reductions in healthcare procedures (see Figure 2 and Figure SI-2a). However, we also document stark differences in the use of oncological procedures between sociodemographic groups, particularly for critical procedures. By the end of 2020, females were almost twice as likely to miss a critical oncological procedure compared to males, and elderly individuals (76 or above) were 14 times less likely to receive critical treatment compared to younger individuals (18–29) (Figure SI-2b).

**Figure 4:**
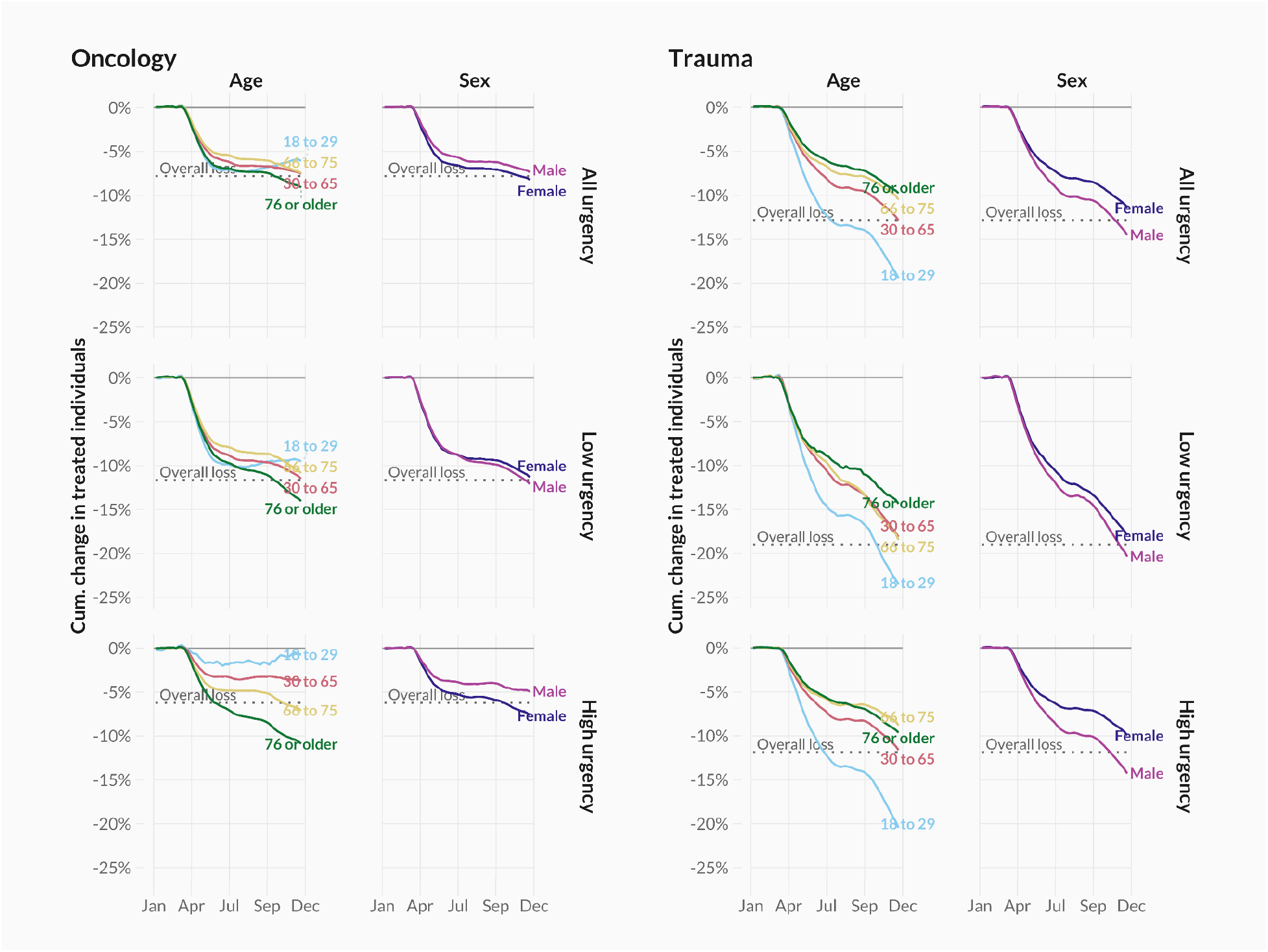
Cumulative age- and sex-adjusted difference between the observed and predicted number of treated individuals in 2020, across urgency types (rows) for a selection of demographic groups (columns), for treatments related to oncology care (left column) and trauma care (right column).

In fact, among 18 to 29 year olds, there is almost no reduction in critical oncological care by the end of 2020.^4^

The findings documented here are robust to a series of empirical robustness checks. These include subsetting the universe of healthcare procedures to i) exclude all diagnostic laboratory tests (Figures SI-4a and SI-4b), and to ii) only consider procedures that included a clinical or emergency room activity (Figures SI-5a and SI-5b). We also re-estimate results using the weekly number of performed healthcare activities as the unit of analysis, rather than the weekly number of individuals receiving care (Figures SI-6a and SI-6b). Across these specifications, we find similar patterns in healthcare declines and inequalities during the pandemic. The accuracy of our estimates rest on the extent to which predictions of healthcare use in 2020 would have mirrored observed healthcare use, had the pandemic not occurred. To gauge the validity of this assumption, we perform a placebo analysis where we set 2019 as the year of interest, and predictions closely mirror observed healthcare use that year (Figures SI-7 and SI-8).

Finally, we assess the possibility that our observed findings are due to selective mortality from COVID. Given that the frail and the elderly were disproportionately likely to use the healthcare system and to die from COVID, mortality due to COVID may have reduced healthcare use that would have otherwise have occurred. In Table SI-1 we display the number of COVID-19 related deaths by demographic characteristics. In line with findings elsewhere, we observe a disproportionate number of deaths amongst the elderly, the poor, and those with a migrant background. However, the number of deaths attributed to COVID accounts for less than 0.2% of the unique recipients of healthcare in 2020, suggesting that mortality selection will have little to no bearing on our substantive results.

## Discussion

The COVID-19 pandemic caused immense strain on healthcare systems as the world battled a novel and ever-evolving disease. In anticipation of COVID hospitalisations and in order to limit further transmission, non-essential care was postponed. In parallel, government interventions were put in place to limit the spread of COVID. In The Netherlands, this meant partial lockdowns from March to May 2020, and again from October to December 2020. Together, these events resulted in dramatic declines to non-COVID healthcare procedures, a pattern that is observed worldwide. We contribute to this literature by analysing howreductions to non-COVID healthcare have been distributed across sociodemographic groups. Our study capitalises on unique full population insurance data from the Dutch universal health coverage system and population register data to document the total decline in healthcare use and to examine how this decline differed by sociodemographic characteristics.

We estimate that by the end of 2020 there were 3 million fewer weekly patients receiving care than would have otherwise, had the pandemic not occurred. This decline was many times larger than the increase in individuals receiving COVID healthcare; during the week of 23rd of March, at the start of the pandemic, there were more than 6,000 additional individuals receiving treatment for COVID across the Dutch healthcare system, while the number of non-COVID related patients decreased by more than 300,000. These numbers illustrate the challenges of adapting a healthcare system to a novel disease, which included increased care requirements^5^, policies to limit transmission to other patients (including partial closures of hospitals), as well as elevated sick leave amongst staff.

The various policies put in place to limit hospitals from being overwhelmed due to COVID hospitalisations should, however, not have affected sociodemographic groups differently. This makes the large social inequalities that we observe in non-urgent procedures particularly concerning. It is possible that historically disadvantaged groups had a lower need for healthcare during 2020. However, it is equally plausible that these groups either could not access the healthcare system during this time of duress or had more difficulty navigating it than their advantaged peers. Although access to healthcare is universal, the Dutch system consists of various types of healthcare providers. Some require patient outreach to utilise, such as finding clinics with capacity for certain procedures and contacting those clinics to sign up for care. More privileged individuals may have experienced fewer barriers to access these types of providers than marginalised groups.

Our findings are especially troubling as the potential for inequality is likely smallest in countries with universal healthcare systems, like The Netherlands. Other healthcare systems, such as the United States, rely heavily on both private insurance and private providers, which introduce inequalities in both the access to and navigation of care. The Netherlands could be viewed as a best case scenario with theoretically equal access, an excellent infrastructure, broadband coverage and an advanced healthcare system. By demonstrating that the pandemic disproportionately impacted health use among marginalised population groups in an otherwise equitable healthcare system, we contribute to a growing body of literature on the unequal nature of the COVID-19 pandemic that goes beyond its direct toll on mortality and disease burden (Bambra et al. 2020; Chen and Krieger 2021; Engzell et al. 2021; Mamelund and Dimka 2021).

Besides differences in access, we also find suggestive evidence for other behavioural mechanisms that may have led to the observed declines in healthcare usage, as well as differences amongst sociodemographic groups. Strong declines in highly urgent trauma-related care, especially amongst young men, likely reflect the effects of government lockdown and subsequent declines in events that might lead to the need for trauma-related care. In the spring of 2020, all public events and large gatherings were prohibited across The Netherlands and in the winter of 2020, a stringent lockdown was implemented, where most non-essential facilities were closed down. Both periods coincide with reductions in highly urgent trauma-related care.

At the same time, we find troubling evidence for declines in healthcare procedures that should not have materialised, including a 6% decline in highly urgent oncological treatments. Although it is reassuring that amongst the young (18-29 year olds) we observe almost no reduction by the year’s end in the number of weekly patients receiving highly urgent oncological care, the observed differences between the old and the young and between females and males are worrying. While pausing some oncological treatments, such as screenings, might have been a necessary decision at the time, there is potential that it may induce excess deaths from cancers in the future (Sharpless 2020). Our findings of large declines in non-urgent oncological care are consistent with findings in The Netherlands, as well as globally, of a pandemic-induced cancer screening backlog (Bakouny et al. 2021; Dinmohamed et al. 2020; Greenwood and Swanton 2021). We already see some possible signs of the downstream consequences of missed cancer screenings in the form of a considerably stronger decline in high urgency oncological care amongst females compared to males, which could be a result of disruptions to the nationwide screenings for breast and cervical cancer during COVID-19 that are unique to females.^6^

Another possible explanation for both the overall declines in non-COVID healthcare as well as the demographic differences in these declines could be hesitance to seek care. Individuals were weary of entering into the healthcare system during the pandemic, for fear of infection (Czeisler et al. 2020; Hafner 2020; Liao et al. 2020; Splinter et al. 2021), making it plausible that at least part of the decline in healthcare use can be explained by individuals not seeking the care they need. Our finding that young individuals were considerably less affected by reductions in care and were more likely to receive non urgent healthcare compared to older individuals is consistent with differing risk assessments regarding COVID infections for different age groups.

The pandemic has affected healthcare through a myriad of processes, ranging from institutional practices that have limited the supply of healthcare, government interventions that have altered the nature of healthcare demand, and more general behavioural changes on healthcare use. We exploit unique data encompassing all individual-level healthcare procedures in The Netherlands linked with rich sociodemographic variables to provide the first complete assessment of declines in healthcare use during the pandemic by sociodemographic characteristics. We find that sociodemographic groups differed considerably in their reduction of healthcare use. These findings are important for policymakers as they continue efforts to make up for delayed care, but also in better understanding the collateral impact of health crises like the COVID-19 pandemic beyond its direct toll on health and well-being.

Further work is necessary to better understand how the differences in healthcare use have materialised. For example, our data does not include information on care provided by general practitioners (GPs), who are often the first point of care and typically provide guidance on whether to send patients to specialist care at a hospital. Studying care practices at the GP level could provide further insights into possible inequities in access, and the extent to which observed declines in healthcare procedures occur as a result of hesitancy from GPs – who may have opted to reduce their referrals to the hospital – or from hesitancy from potential patients – who may have opted not to seek care. Similarly, we only measure use of the healthcare system and not objective need. Complementing observed use with need would provide further pointers to disentangle the extent to which avoidance of care has driven overall declines and differences amongst sociodemographic groups.

Our study serves as a reminder that the health consequences of the COVID-19 pandemic span beyond mortality and long COVID, to include a profound impact on non-COVID related healthcare use. Importantly, we show that the burden of declines in healthcare use has not been distributed equally. Although our study takes place in the Dutch context, where the universal healthcare provision strives to minimise healthcare inequalities, we still observe strong inequities in reduced healthcare. These disparities are likely considerably smaller than in other contexts, where baseline access to healthcare is systemically unequal, such as in the United States. As policymakers and healthcare professionals strive to catch up on missed care, it is critical to understand that targeted efforts to reach historically marginalised and disadvantaged population groups is of utmost importance.

## Methods

We use Dutch registry data, accessed through the Remote Access environment hosted by the Centraal Bureau voor de Statistiek (CBS 2021). The registry contains individual-level records of all persons residing in The Netherlands each year, including detailed information on their social, demographic, and economic background (see Table SI-2 for descriptive statistics of the population under study). In addition, the registry contains information on every single health expenditure that was paid for by the universal health insurance system in The Netherlands. This includes all care that has been performed in hospital and was covered by the Dutch universal healthcare system. The dataset does not include data from general practitioners, as information on general practitioner care was not available.^7^

Every activity covered by the universal health care system is logged and assigned various classifications, including the Diagnosis-Treatment-Classifications (DTCs) that an activity falls under. The DTC reflects a substantive diagnosis and treatment plan and allows us to assign activities to medical groups like Oncology or Trauma. Each activity is time-stamped at the daily level and can be assigned an urgency level through the classification of DTC’s by the Dutch Healthcare Authority (NZa). There are seven levels of urgency, ranging from extremely urgent (with a planning window of less than 24 hours) to non-urgent (with a planning window greater than 3 months). We assign procedures to one of three levels of urgency, with ‘high urgency’ procedures having a planning horizon of less than a week, ‘middle urgency’ procedures having a planning horizon of between one week and less than two months, and ‘low’ urgency procedures a planning horizon of two months or more. Yearly counts of the number of activities can be found in Table SI-3.

In our analytical approach, we calculate the number of unique individuals for each age and sex that had at least one activity performed in a given week for the period 2017-2020. This generates a set of weekly counts of unique individuals that were treated for every combination of age and sex. Note that if an individual seeks treatment in two weeks in a given year, she is counted twice. We calculate these time series for various subsets of activities and demographics. We then age-sex standardise the number of individuals treated per week to the full population in 2018, to ensure that our findings are not driven by differences in age and sex composition between groups and over time.

To generate estimates of the expected number of individuals that would have been treated in the period between March and December of 2020, we estimate a linear model using the weekly counts of individuals as the outcome based on data up until including February 2020. Our model includes week fixed effects, the number of days in the week (to address differences in the first and last weeks of the year) and the number of holiday days. We also include year fixed effects. We use this model to make predictions for every week between March and December 2020.^8^ We estimate separate models for every demographic and/or activity subset, for example when considering the number of individuals with a non-Western background that made use of low urgency activities (see Figure SI-9).

Finally, we use our estimates of the weekly number of individuals receiving care in the system and compare these with the observed number of weekly individuals. This analytical approach is illustrated in Figure SI-8. We show the differences between the expected and realised number of treated individuals as a weekly difference, as well as a cumulative sum over successive weeks. Note that we exclude all healthcare activities that are associated with a COVID-19 infection. We identify these activities as those DTC’s that were assigned a COVID-19 ICD-10 code.

## Data Availability

All data produced in the present study are based on non-public data available at the Centraal Bureau voor de Statistiek pending institutional approval.

## Data Availability

All results presented here are calculated from non-public registry data from Centraal Bureau voor de Statistiek (CBS), accessed through the Remote Access environment. CBS was not involved in the calculation of any of the results presented here. While the data are not publicly available, academic institutions can apply for access through the CBS.

All code underlying our analyses are available at: https://github.com/MarkDVerhagen/Dutch_healthcare_inequalities_COVID19.

## Supplementary Information

**Table SI-1:** COVID-19 related mortality by sociodemographic characteristics in 2020. *Note*: counts by age group have been rounded to multiples of ten given small sample sizes. As a result, the total count across age groups is higher than the overall total reported in the first row.

|  |  | Population | Covid-19 deaths (prop) |
| --- | --- | --- | --- |
| Total (N) |  | 14,071,420 | 20,125 |
| Gender (%) | Women | 0.51 | 10,755 (0.53) |
|  | Men | 0.49 | 9,370 (0.47) |
| Age Group (%) | 18-29 | 0.19 | 10 (0.0) |
|  | 30-65 | 0.58 | 1,280 (0.06) |
|  | 66-75 | 0.13 | 3,330 (0.17) |
|  | 76+ | 0.09 | 15,510 (0.77) |
| Migrant background (%) | Native | 0.76 | 16,795 (0.83) |
|  | Migrants | 0.24 | 3,330 (0.17) |
| Poverty (%) | Not poor | 0.91 | 16,980 (0.84) |
|  | Poor | 0.09 | 3,145 (0.16) |

**Table SI-2:** Average number of individuals treated per week between 2017 and 2020 (based on 52 weeks) and by demographic group. *Note*: An individual is counted as having received treatment in a given week if they underwent at least one health activity that week. If an individual received at least one health activity per week across two weeks, they will be counted twice.

|  |  | 2017 | 2018 | 2019 | 2020 |
| --- | --- | --- | --- | --- | --- |
| Average patients per week (N) |  | 590,769 | 620,059 | 627,573 | 570,884 |
| Gender (%) | Women | 0.56 | 0.56 | 0.56 | 0.55 |
|  | Men | 0.44 | 0.44 | 0.44 | 0.45 |
| Age Group (%) | 18-29 | 0.08 | 0.08 | 0.08 | 0.09 |
|  | 30-65 | 0.51 | 0.51 | 0.50 | 0.50 |
|  | 66-75 | 0.22 | 0.23 | 0.23 | 0.23 |
|  | 76+ | 0.17 | 0.17 | 0.18 | 0.18 |
| Migrant background (%) | Native | 0.81 | 0.80 | 0.80 | 0.80 |
|  | Migrants | 0.19 | 0.20 | 0.20 | 0.20 |
| Poverty (%) | Not poor | 0.88 | 0.88 | 0.88 | 0.89 |
|  | Poor | 0.12 | 0.12 | 0.12 | 0.11 |

**Table SI-3:** Total count of individuals treated per week between 2017 and 2020 by urgency type. *Note*: An individual is counted as having received treatment in a given week at a given urgency level if they underwent at least one health activity with that urgency level that week. If an individual received both a high urgency and a low urgency treatment within a given week, they will appear in both urgency counts.

|  | 2017 | 2018 | 2019 | 2020 |
| --- | --- | --- | --- | --- |
| Total | 30,719,977 | 32,243,093 | 32,633,772 | 29,685,982 |
| High urgency | 6,263,624 | 6,402,855 | 6,405,455 | 5,838,573 |
| Middle urgency | 7,822,592 | 8,387,678 | 8,450,859 | 7,914,615 |
| Low urgency | 14,150,766 | 14,809,763 | 14,990,769 | 13,322,366 |
| No urgency | 4,450,758 | 4,806,731 | 4,978,988 | 4,601,216 |

**Table SI-4:** Total count of treatment activities between 2017 and 2020 and by demographic group

|  |  | 2017 | 2018 | 2019 | 2020 |
| --- | --- | --- | --- | --- | --- |
| Total (N) |  | 90,207,501 | 91,472,813 | 92,208,940 | 81,108,838 |
| Gender (%) | Women | 0.55 | 0.55 | 0.55 | 0.54 |
|  | Men | 0.45 | 0.45 | 0.45 | 0.46 |
| Age Group (%) | 18-29 | 0.07 | 0.07 | 0.07 | 0.07 |
|  | 30-65 | 0.47 | 0.46 | 0.46 | 0.46 |
|  | 66-75 | 0.22 | 0.23 | 0.24 | 0.24 |
|  | 76+ | 0.22 | 0.22 | 0.23 | 0.23 |
| Migrant background (%) | Native | 0.82 | 0.81 | 0.81 | 0.81 |
|  | Migrants | 0.18 | 0.19 | 0.19 | 0.19 |
| Poverty (%) | Not poor | 0.86 | 0.86 | 0.87 | 0.87 |
|  | Poor | 0.14 | 0.14 | 0.13 | 0.13 |

**Table SI-5:** Total count of treatment activities between 2017 and 2020 and by urgency type

**Table SI-4:** Total count of treatment activities between 2017 and 2020 and by demographic group
|  | 2017 | 2018 | 2019 | 2020 |
| --- | --- | --- | --- | --- |
| Total | 90,207,501 | 91,472,813 | 92,208,940 | 81,108,838 |
| High urgency | 30,937,027 | 30,964,590 | 31,137,706 | 27,608,948 |
| Middle urgency | 17,641,062 | 18,441,306 | 18,505,271 | 16,918,351 |
| Low urgency | 28,914,454 | 29,391,695 | 29,486,959 | 24,899,905 |
| No urgency | 12,714,958 | 12,675,222 | 13,079,004 | 11,681,634 |

**Table SI-6:**
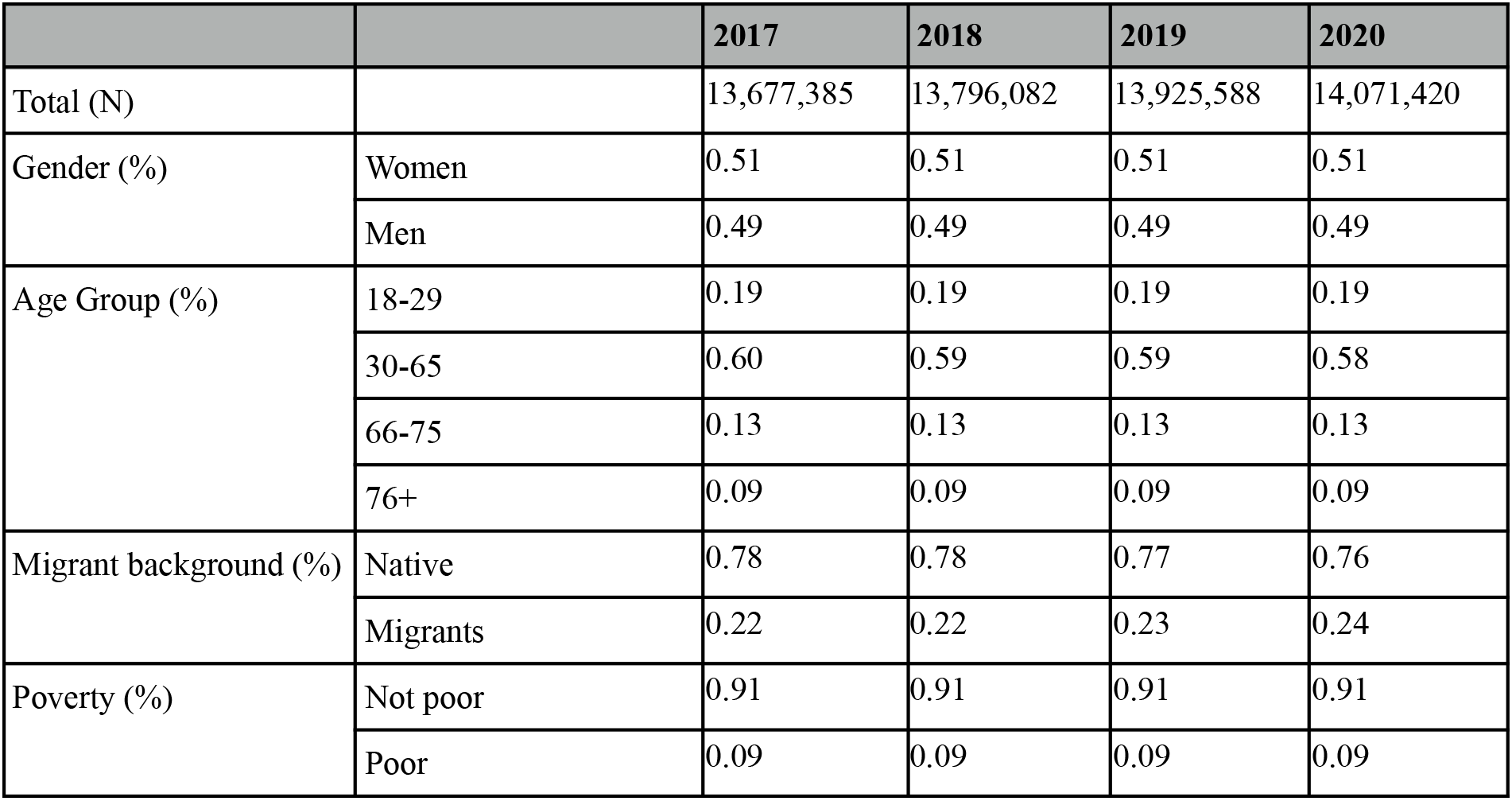
Population composition The Netherlands between 2017 and 2020.

**Table SI-7:**
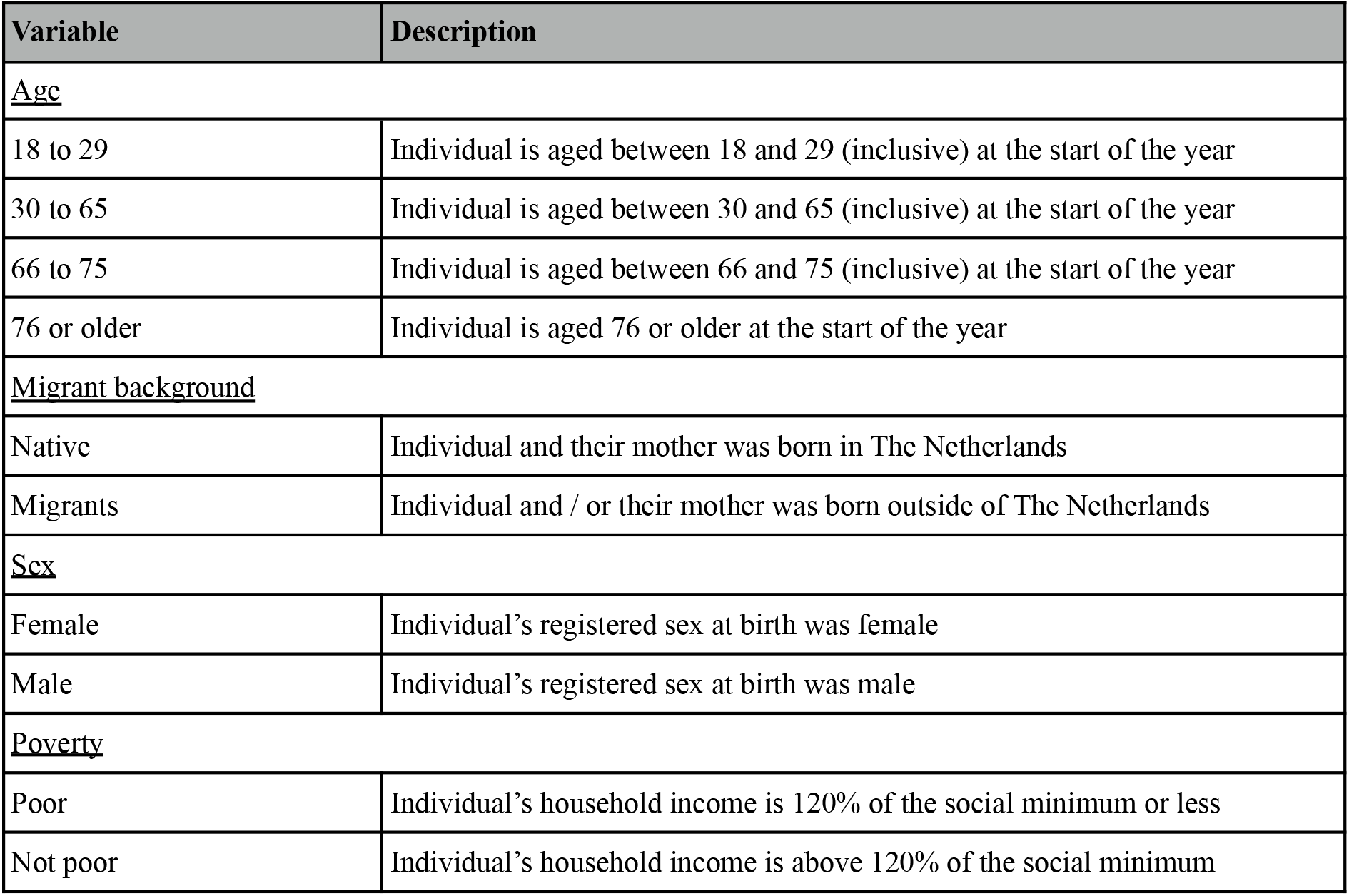
Sociodemographic variables included in the data.

**Figure SI-1:**
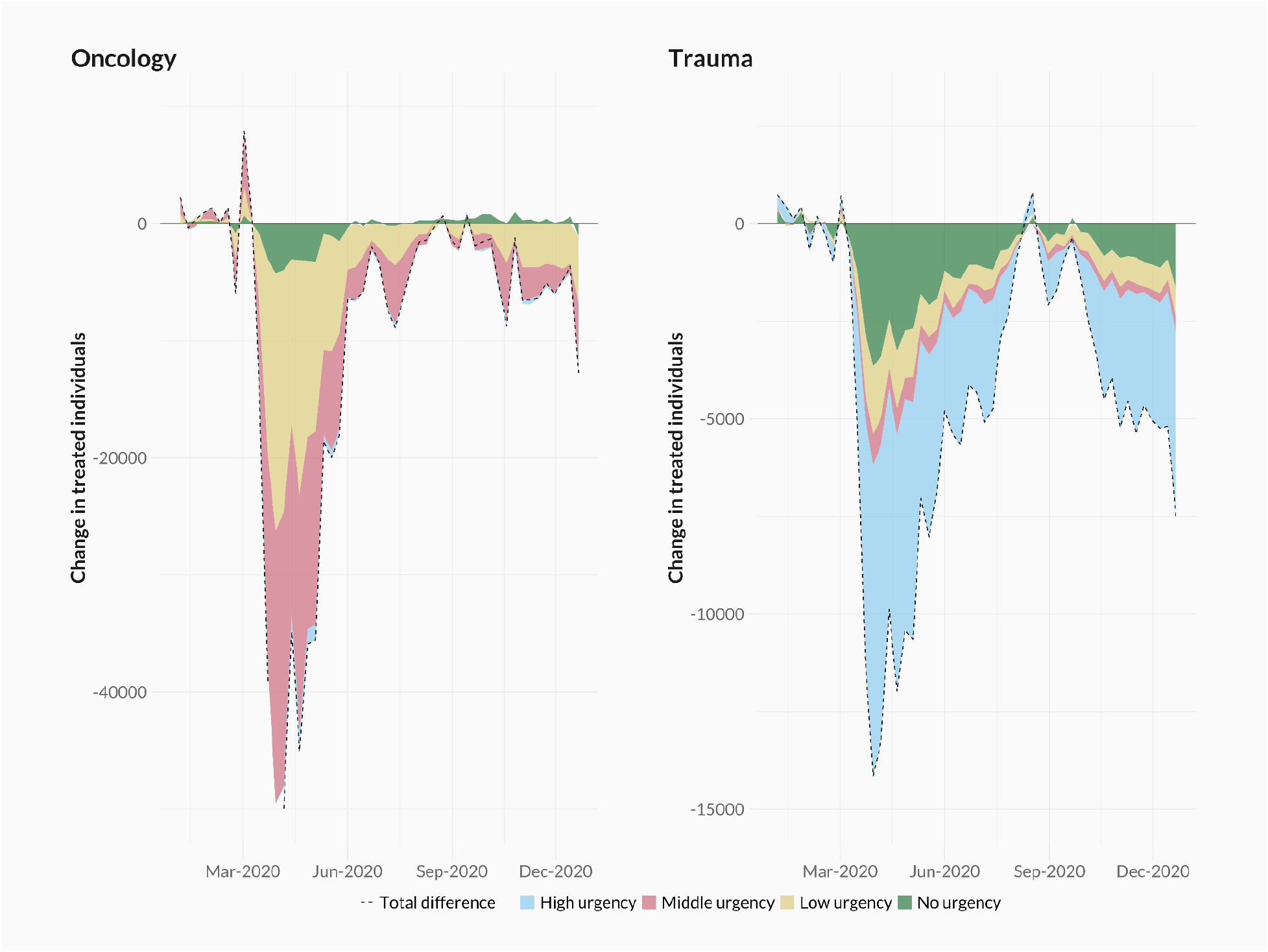
Difference between the observed and predicted number of treated individuals per week in 2020, for oncological care (left) and trauma care (right). Colours differentiate between urgency types (high, middle, low, and no urgency).

**Figure SI-2a:**
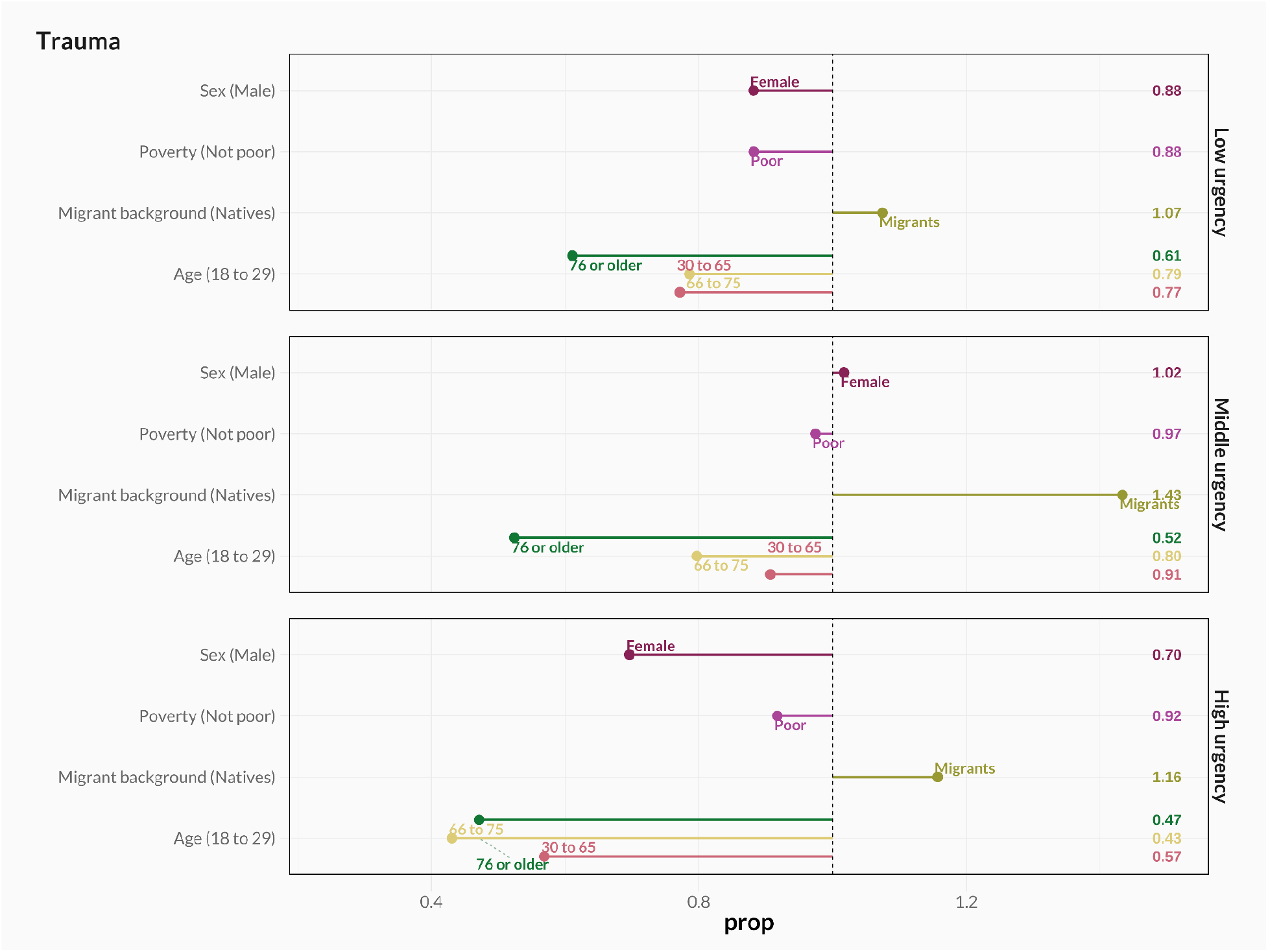
Ratio of the cumulative loss across 2020 for each demographic group relative to a benchmark group (in brackets) for trauma-related healthcare.

**Figure SI-2b:**
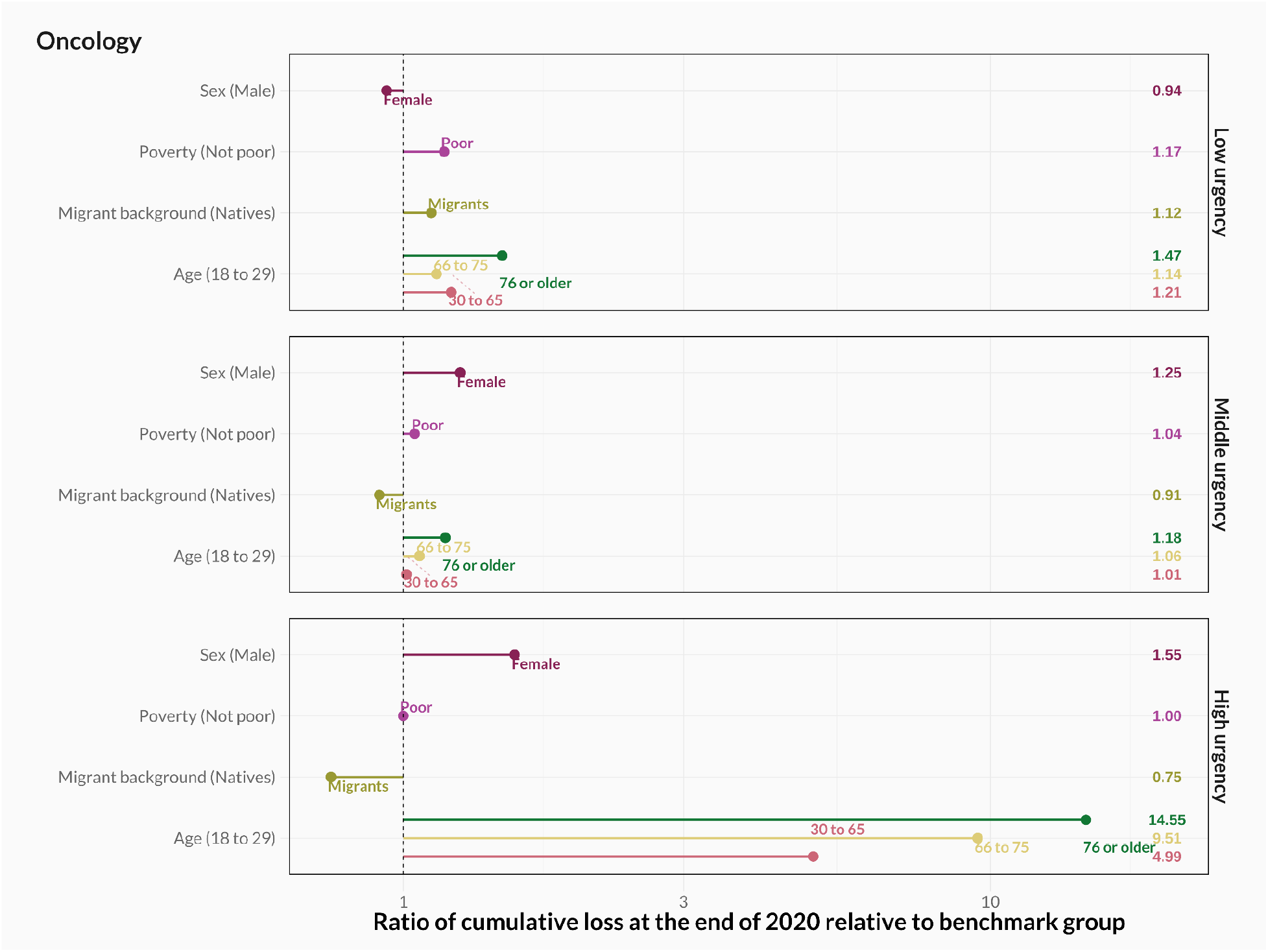
Ratio of the cumulative loss across 2020 for each demographic group relative to a benchmark group (in brackets) for oncological healthcare.

**Figure SI-2a:**
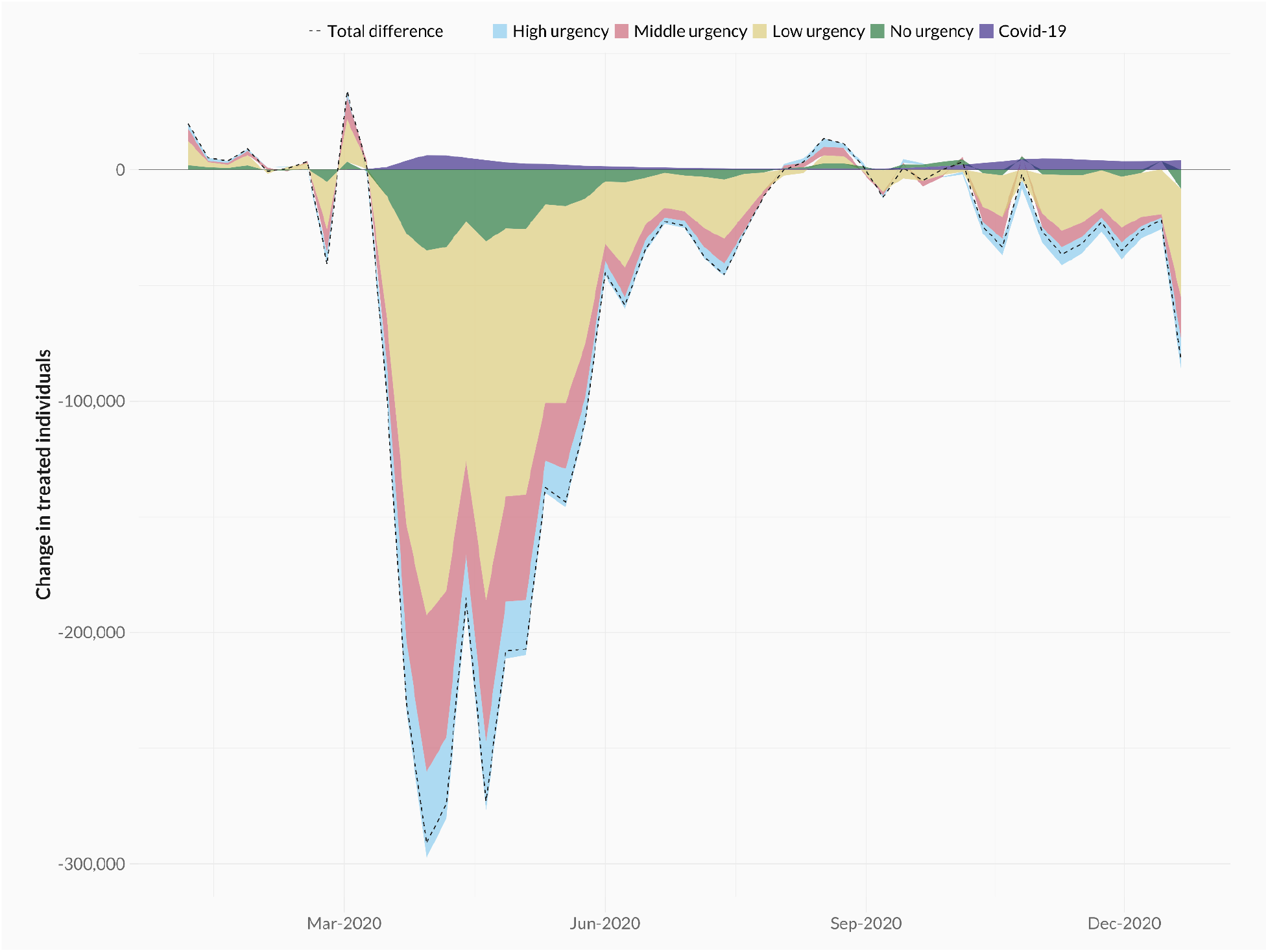
Difference between the observed and predicted number of treated individuals per week in 2020, when only including individuals receiving healthcare procedures involving outpatient visits, clinical activities, ER activities and diagnostics but excluding activities related to laboratory medicine. Colours differentiate between urgency types (high, middle, low, and no urgency).

**Figure SI-2b:**
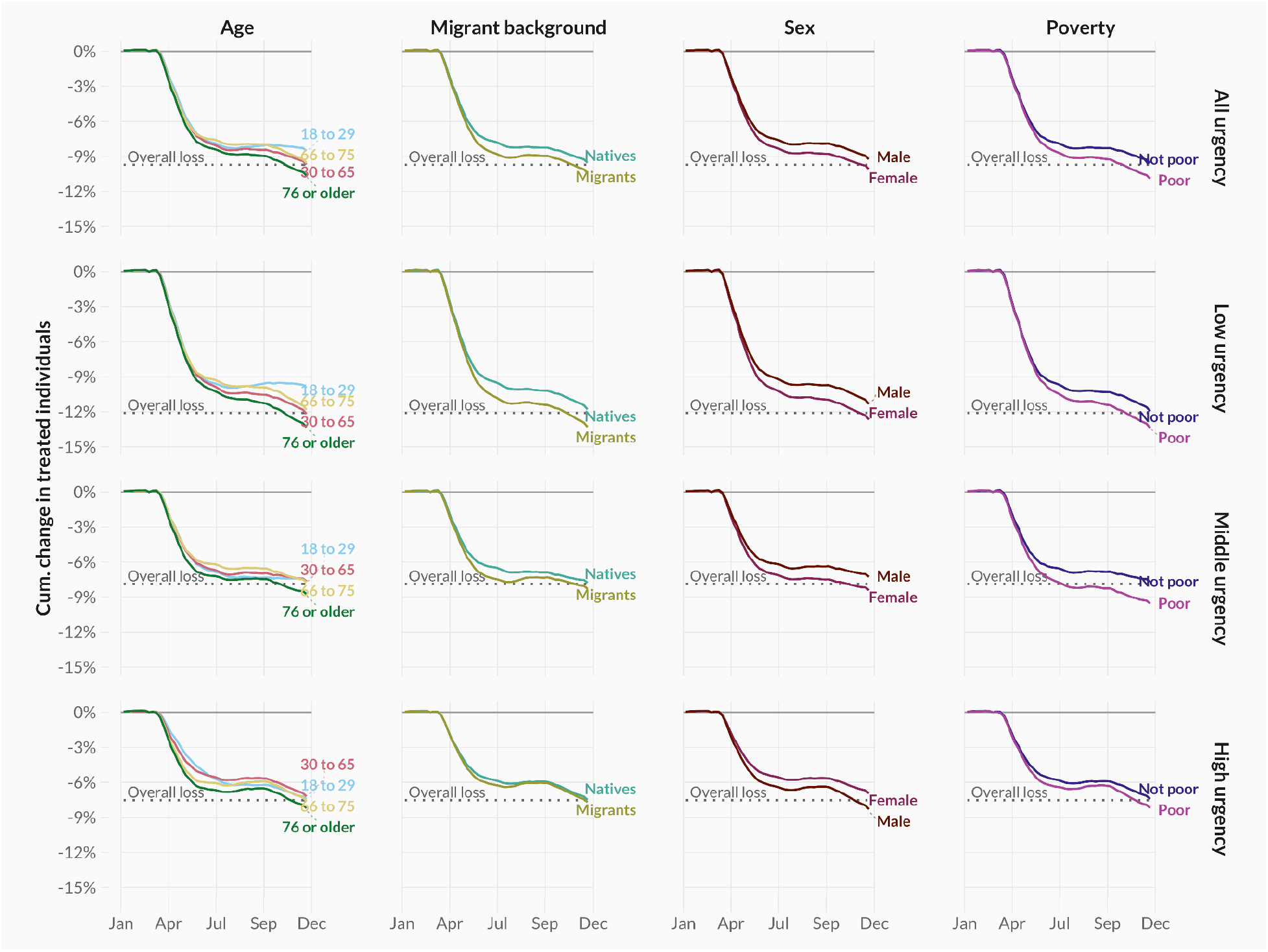
Cumulative age- and sex-adjusted difference between the observed and predicted number of treated individuals in 2020 when only including individuals receiving healthcare procedures involving outpatient visits, clinical activities, ER activities and diagnostics but excluding activities related to laboratory medicine, across urgency types (rows) and demographic groups (columns).

**Figure SI-3a:**
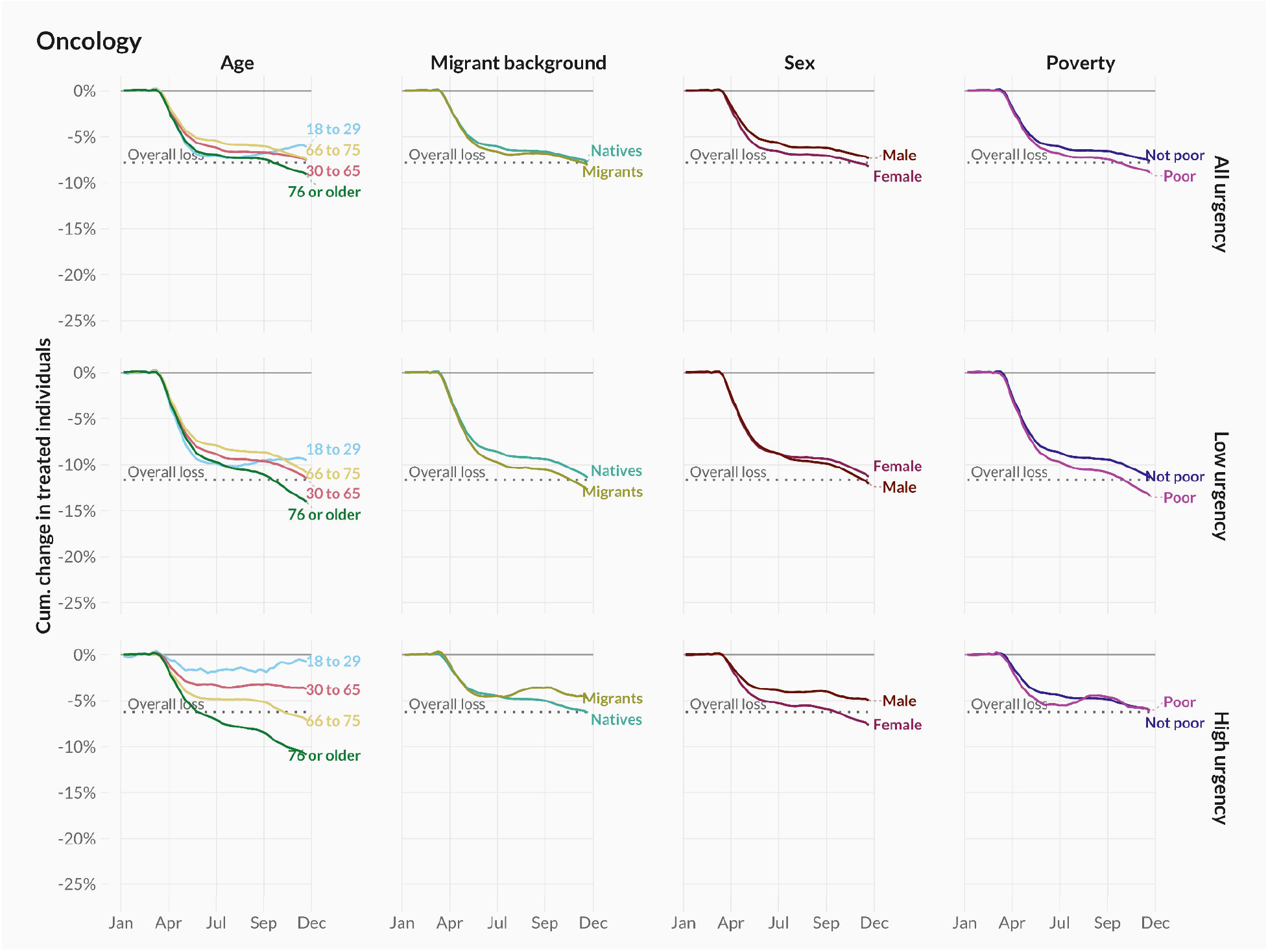
Cumulative age- and sex-adjusted difference between the observed and predicted number of treated individuals in 2020, across urgency types (rows) for all demographic groups (columns), for treatments related to oncology care.

**Figure SI-3b:**
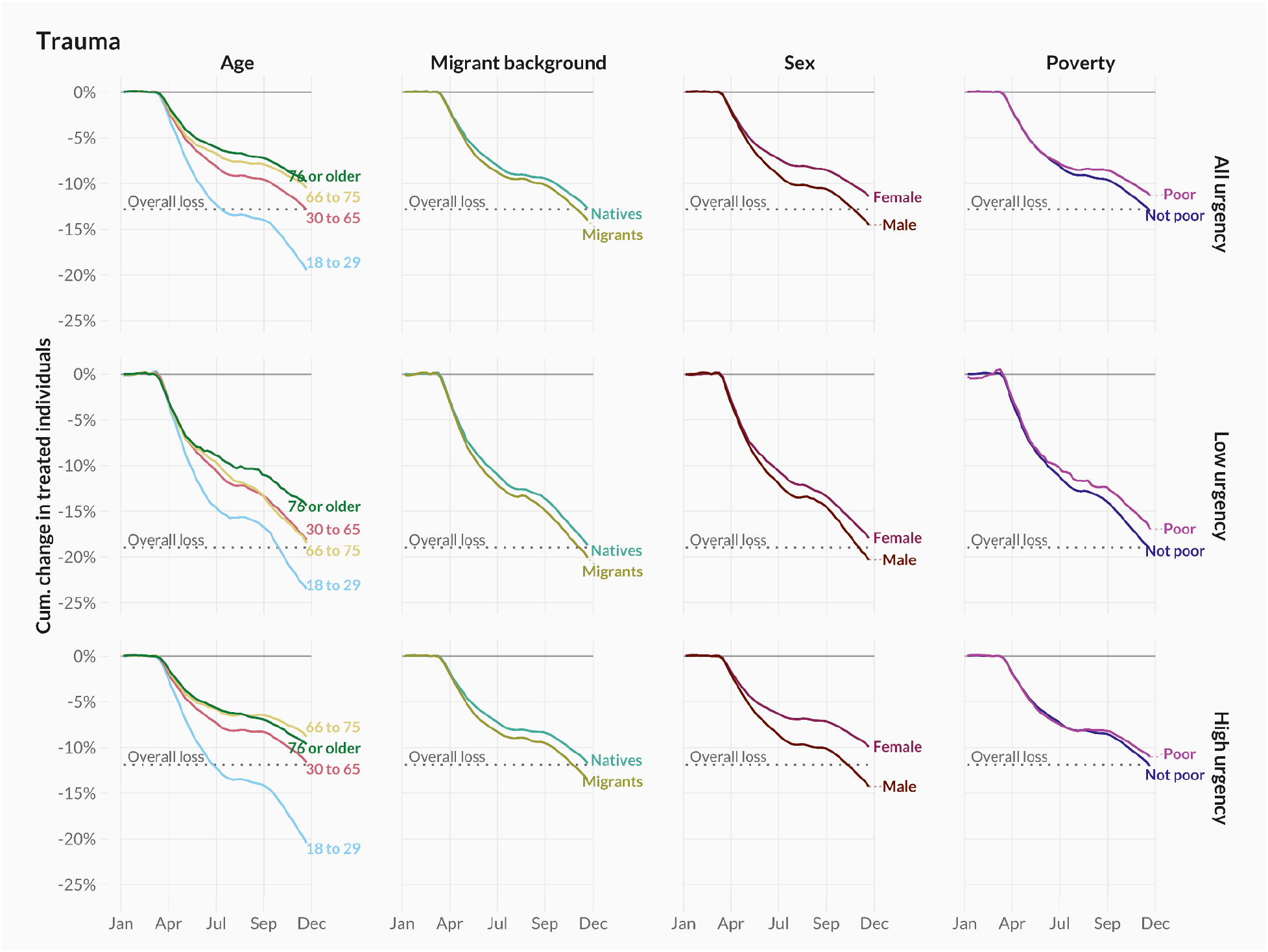
Cumulative age- and sex-adjusted difference between the observed and predicted number of treated individuals in 2020, across urgency types (rows) for all demographic groups (columns), for treatments related to trauma care.

**Figure SI-4a:**
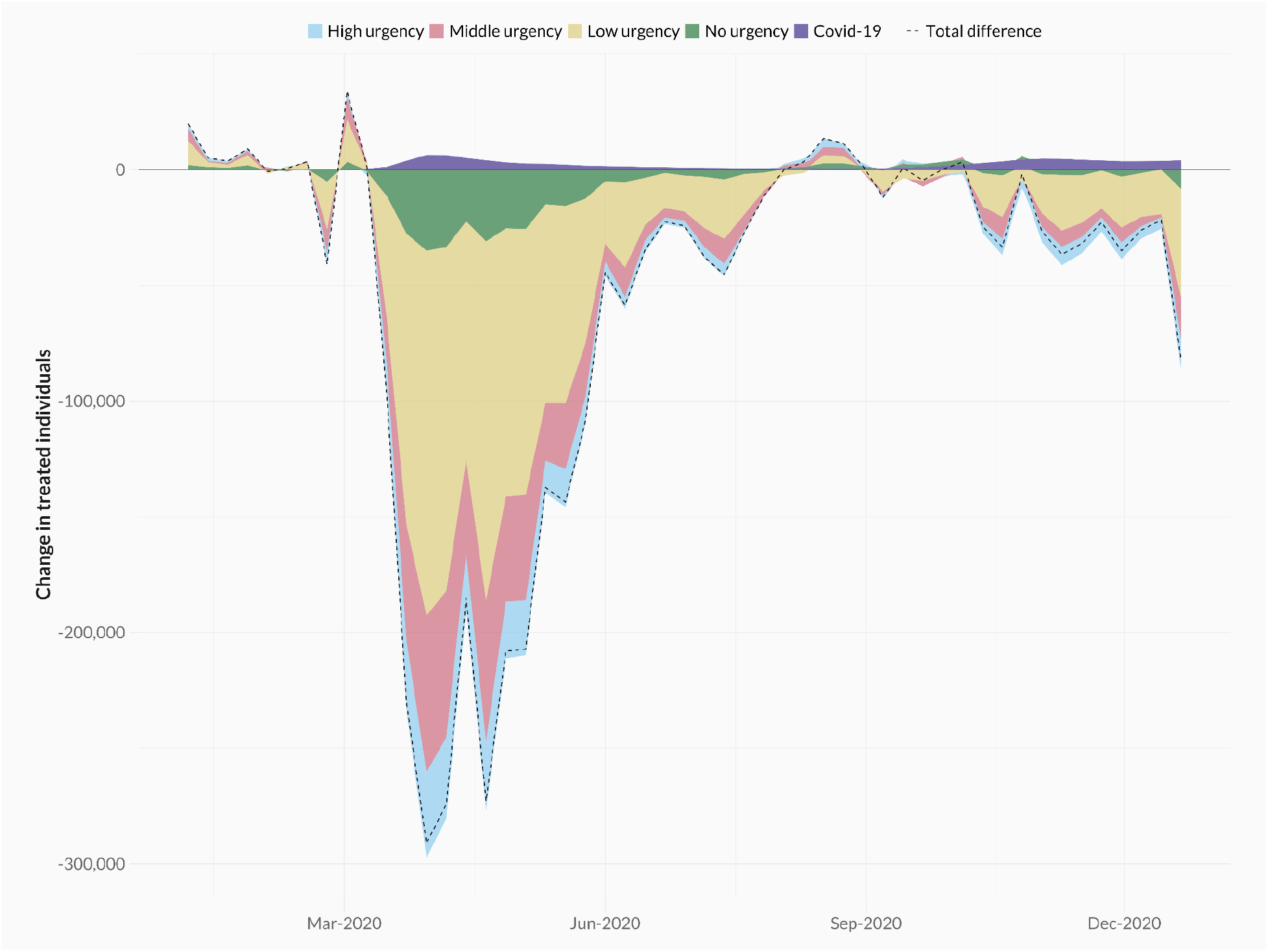
Difference between the observed and predicted number of treated individuals per week in 2020, when excluding all diagnostic laboratory treatments. Colours differentiate between urgency types (high, middle, low, and no urgency).

**Figure SI-4b:**
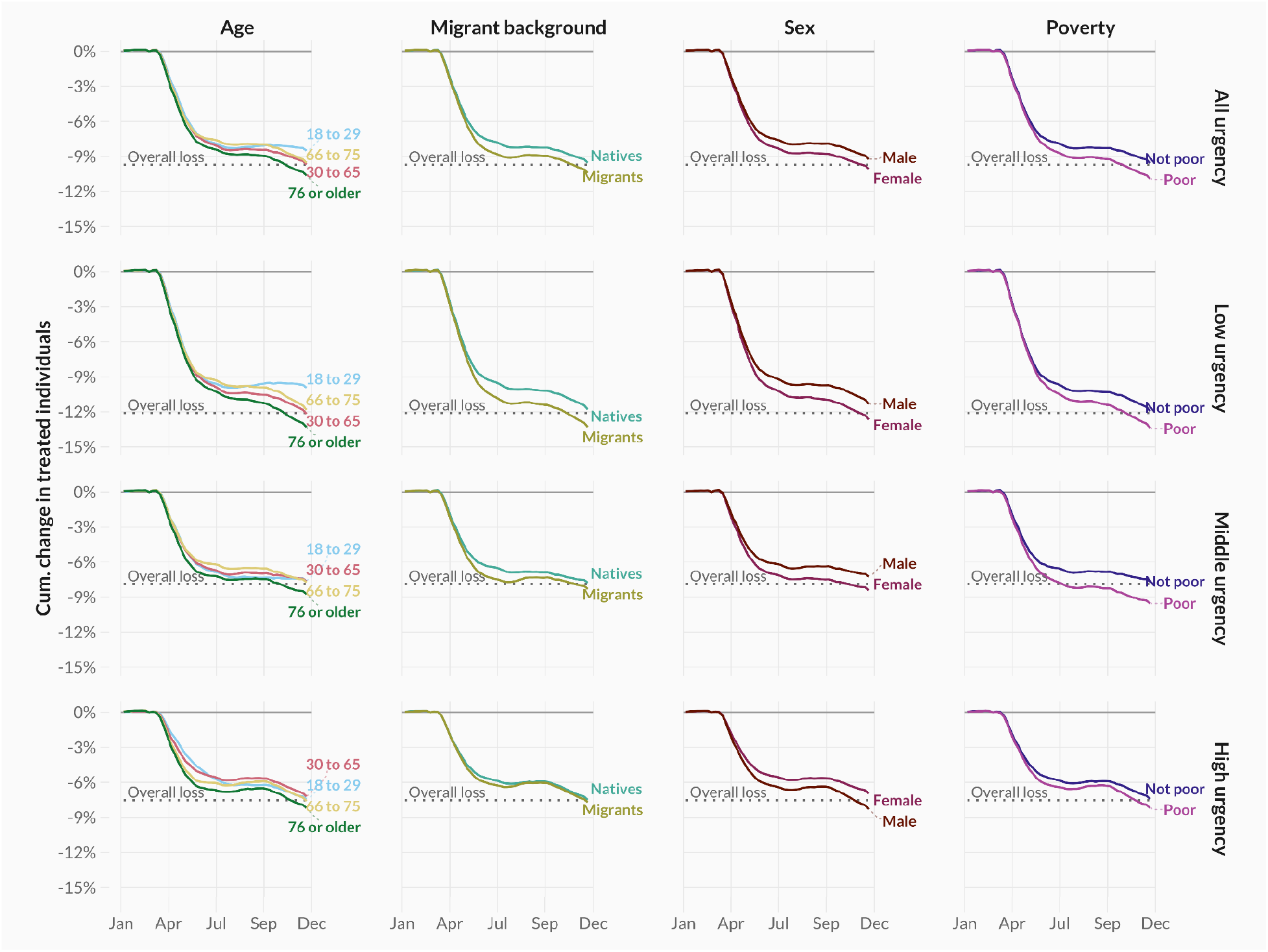
Cumulative age- and sex-adjusted difference between the observed and predicted number of treated individuals in 2020, across urgency types (rows) and demographic groups (columns), when excluding all diagnostic laboratory treatments.

**Figure SI-5a:**
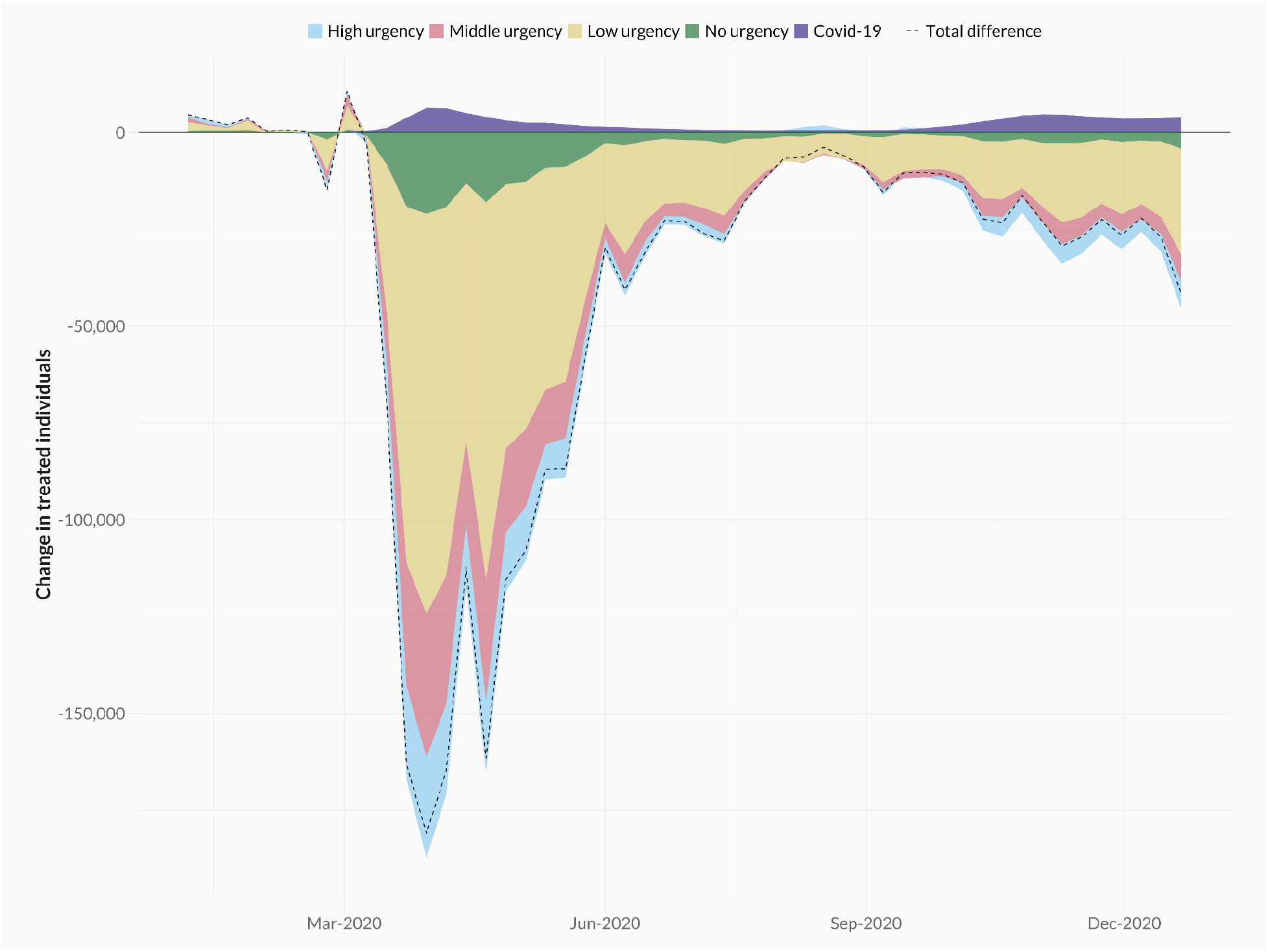
Difference between the observed and predicted number of treated individuals per week in 2020, when only including individuals receiving healthcare procedures involving clinical and / or ER activities. Colours differentiate between urgency types (high, middle, low, and no urgency).

**Figure SI-5b:**
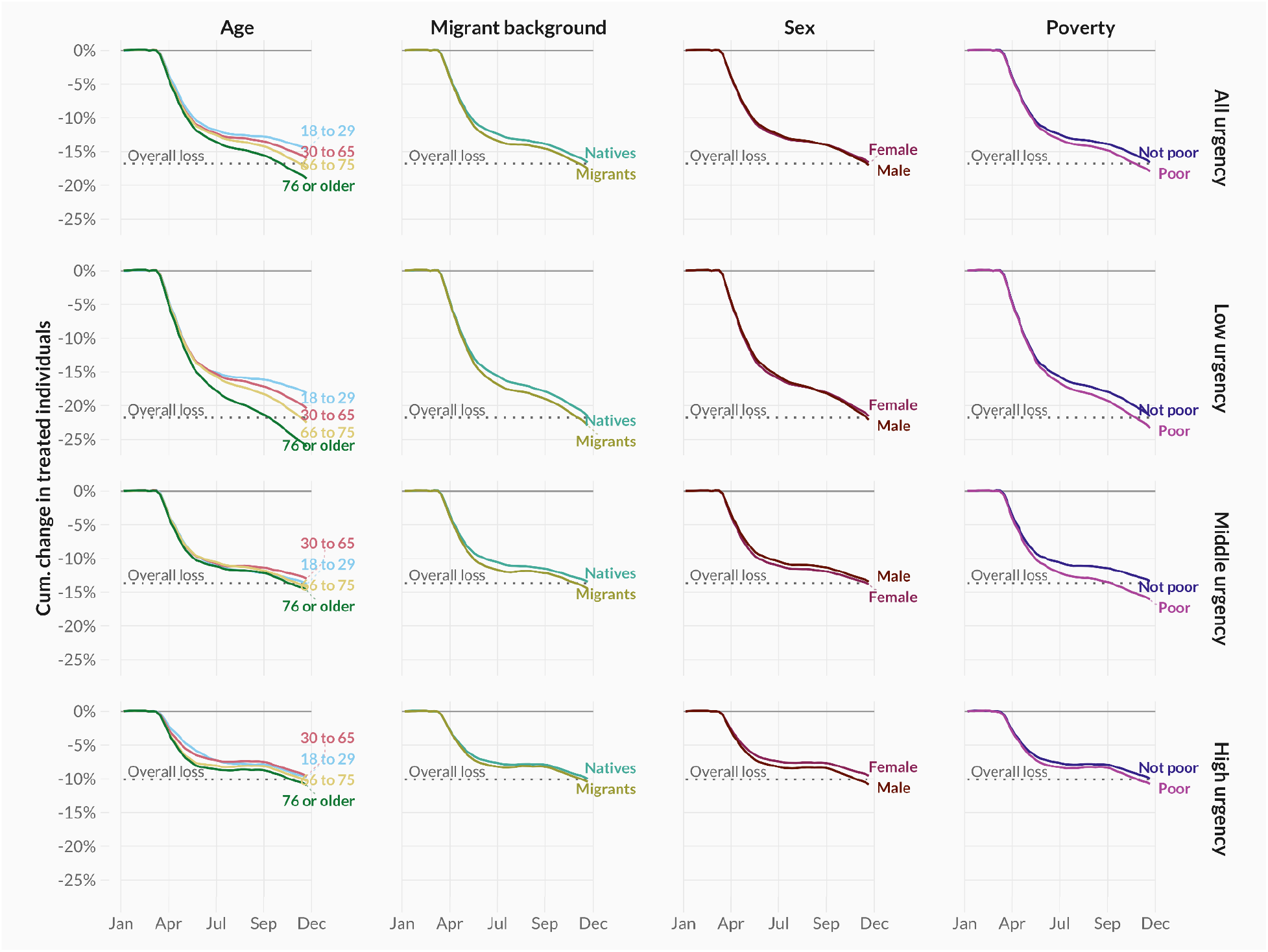
Cumulative age- and sex-adjusted difference between the observed and predicted number of treated individuals in 2020 when only including individuals receiving healthcare procedures involving clinical and / or ER activities, across urgency types (rows) and demographic groups (columns).

**Figure SI-6a:**
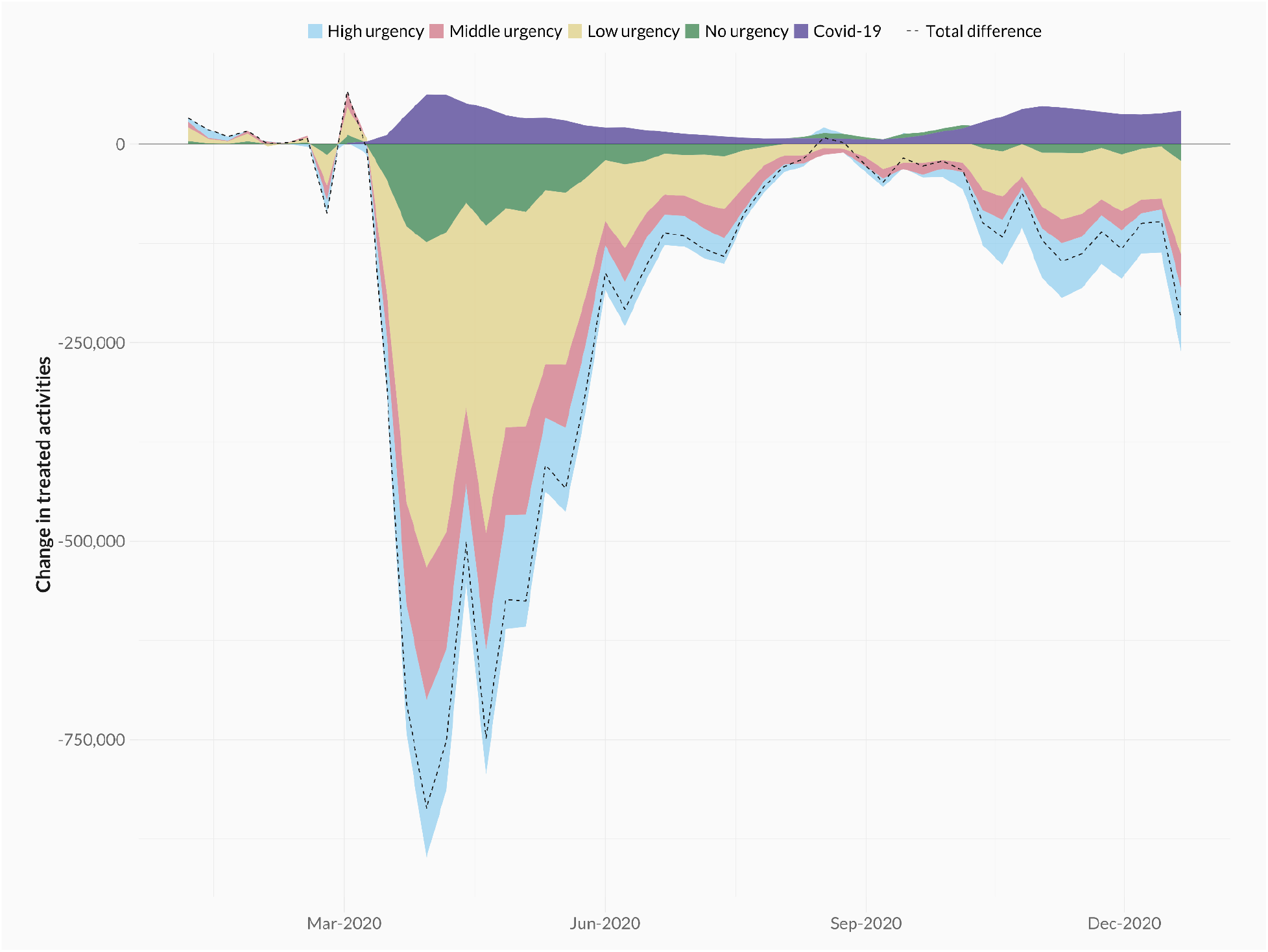
Difference between the observed and predicted number of healthcare activities per week in 2020. Colours differentiate between urgency types (high, middle, low, and no urgency).

**Figure SI-6b:**
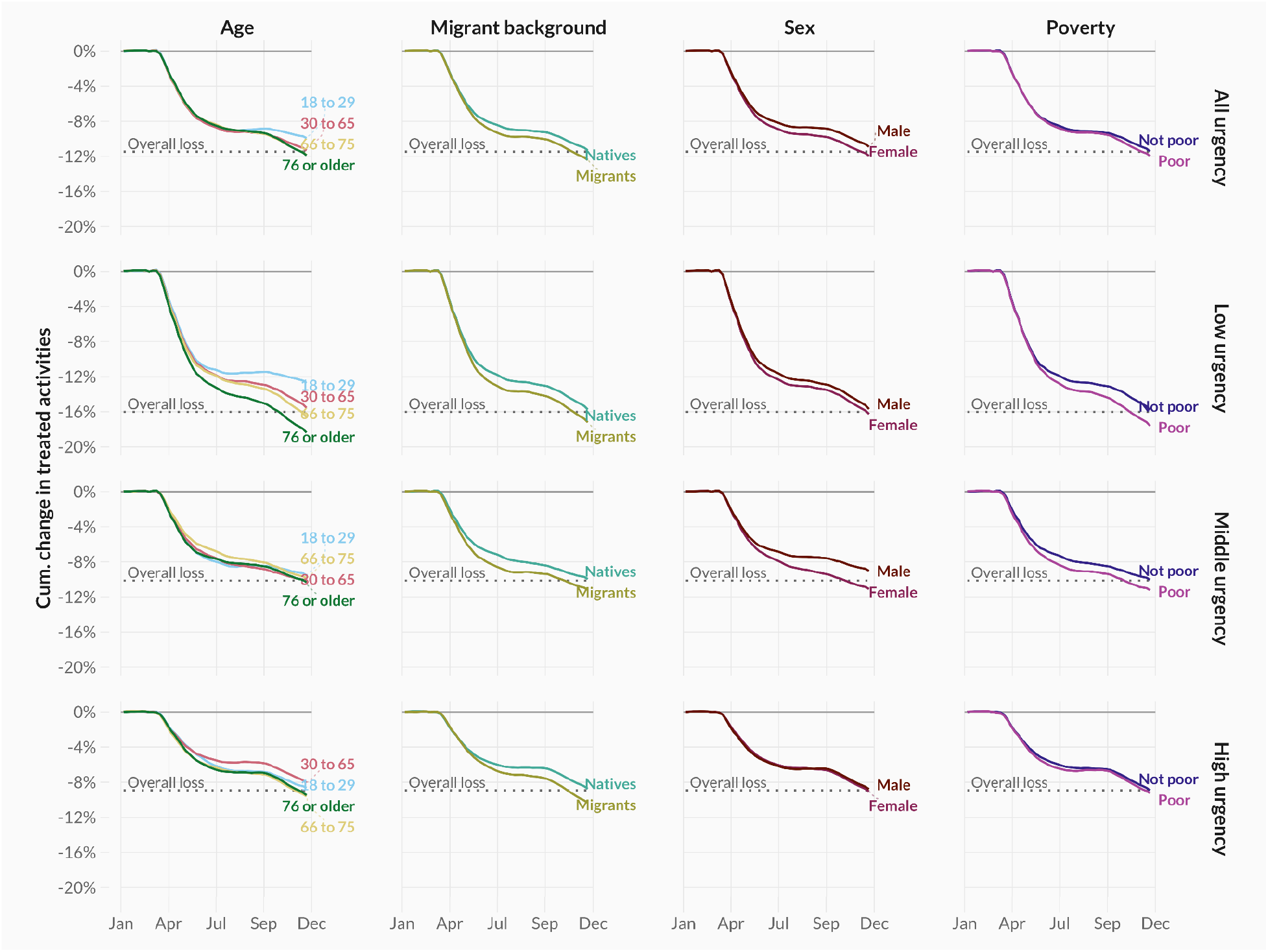
Cumulative age- and sex-adjusted difference between the observed and predicted number of treated individuals in 2020 when considering unique activities for treatments related to oncology care (left column) and trauma care (right column), across urgency types (rows) and demographic groups (columns).

**Figure SI-7:**
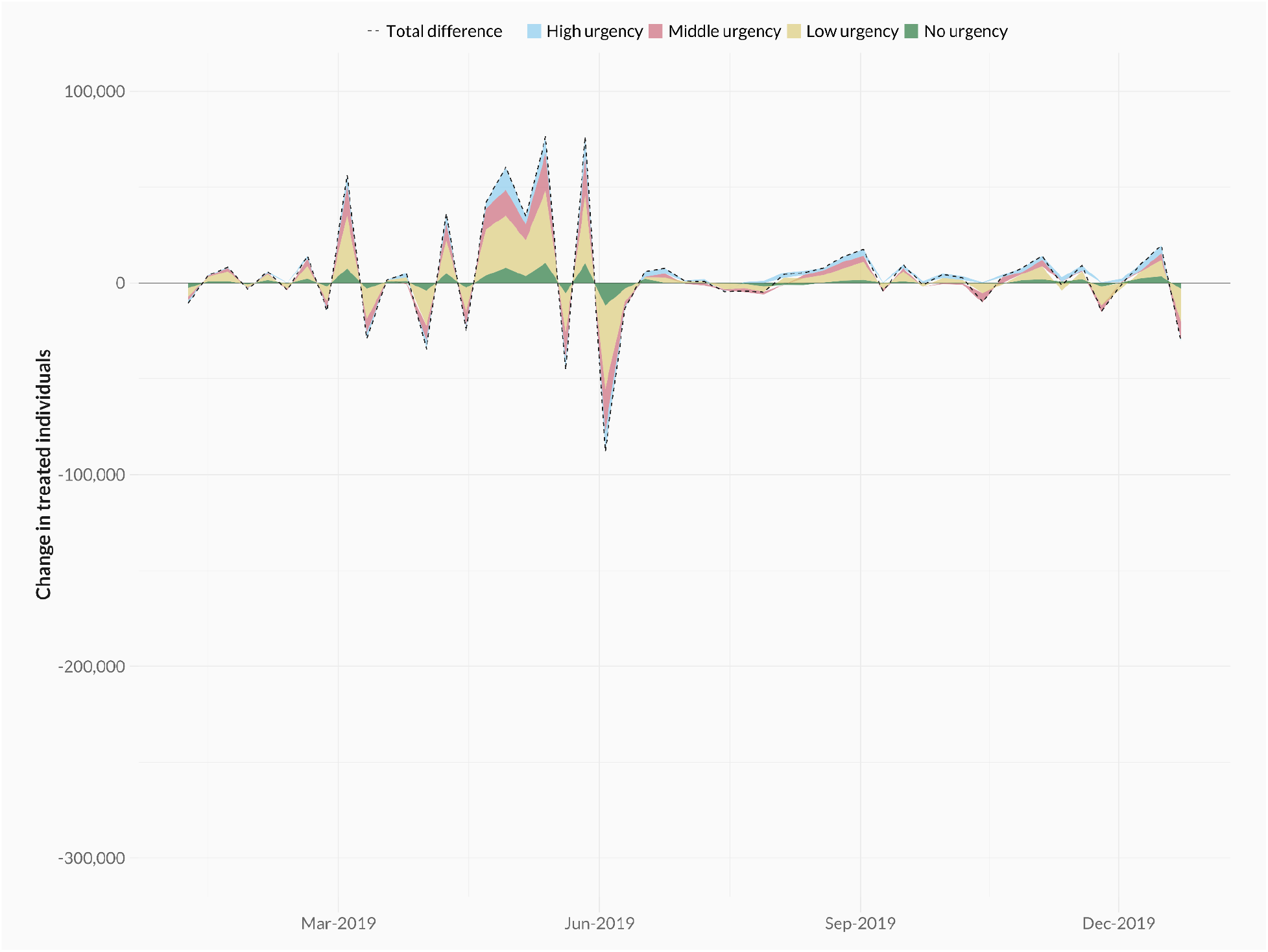
Difference between the observed and predicted number of treated individuals per week in 2019. Colours differentiate between urgency types (high, middle, low, and no urgency).

**Figure SI-8:**
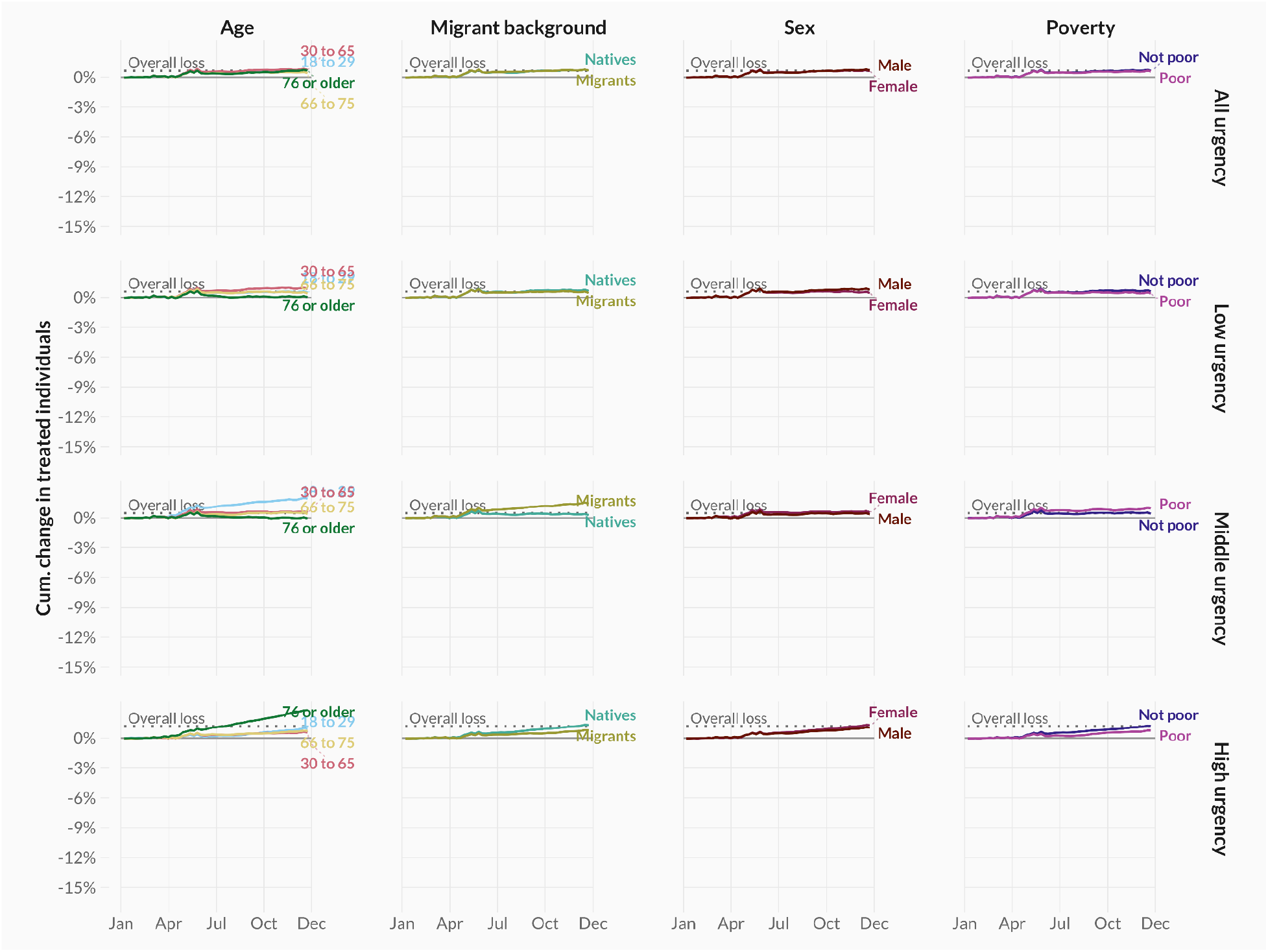
Cumulative age- and sex-adjusted difference in observed versus expected healthcare users by urgency (rows) and demographic groups (columns) in 2019.

**Figure SI-9:**
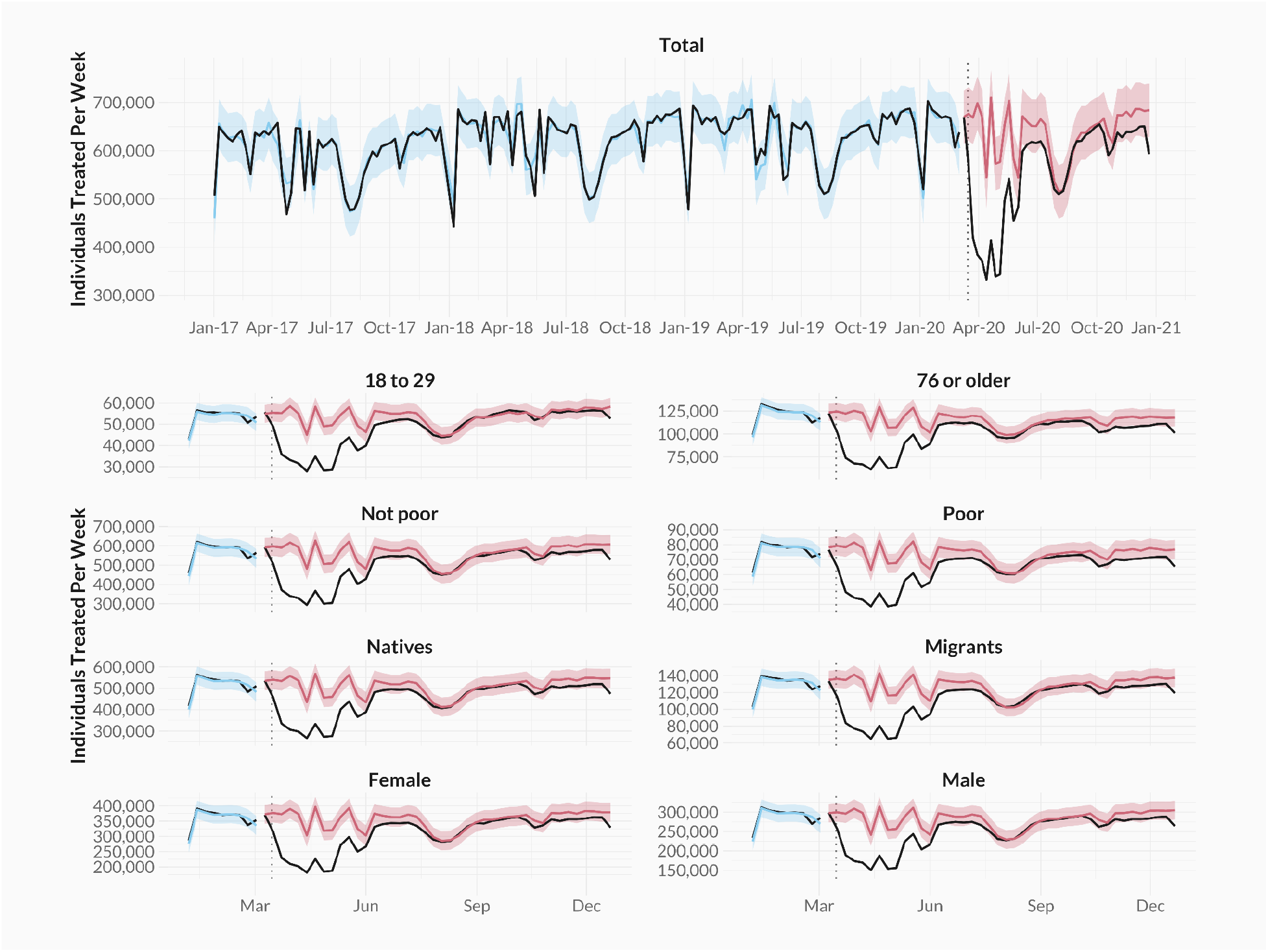
Predicted and observed number of weekly patients, 2017 to 2020. Predicted number of treated individuals per week in blue and red, observed number of treated individuals in black. Colours differentiate between the in-sample (blue) and out-of-sample (red) period.

## Footnotes

1 We consider the weekly number of patients as the number of unique individuals having received at least one healthcare activity during that week. We also consider raw counts of activities as a robustness check further below.

2 An example of highly urgent trauma care is “surgery for an extra-articular trauma to an extremity” (NZa product code 199299069), which was one of the most frequently occurring treatment types in 2019, besides general trauma-related activities (without additional specification).

3 An example of highly urgent oncological care is “hospitalisations due to a malignancy in the brain” (NZa product code 29799070), which was the most frequently occurring treatment type in 2019 besides general post-surgery care.

4 Note that other surprising patterns not discussed in detail here include some recovery in high urgent oncological procedures amongst the migrant population in The Netherlands (Figures SI-3a and SI-3b).

5 The average COVID hospital stay was 8.2 days, compared to 5.9 days for a typical non-COVID procedure involving a hospital stay.

6 In The Netherlands, screenings for breast cancer were stopped at the beginning of the pandemic and only recommenced in an altered form in July 2020, relying more on self testing rather than screenings performed by general practitioners. Other nationwide screenings experienced similar disruptions (for a full account of disruptions during the pandemic, see https://magazines.rivm.nl/2021/04/de-kracht-van-verbinding-2020/corona-en-de-bevolkingsonderzoeken).

7 Universal healthcare covers primary health procedures and excludes, for example, dental care or physiotherapy.

8 While we predict healthcare use for all weeks between March and December, 2020, we exclude weeks 52 and 53 from the plots since those weeks include an irregular number of days.

## Notes

### Competing Interest Statement

The authors have declared no competing interest.

### Funding Statement

We acknowledge research funding and support from the Leverhulme Trust Large Centre Grant (A.F., A.M.T., M.D.V), ZonMW grant 10430252210004 (A.F., A.M.T., M.D.V.), Nuffield College (A.F., A.M.T., M.D.V.) and European Research Council grant ERC-2021-CoG-101002587 (A.M.T.).

### Author Declarations

The Centraal Bureau voor de Statistiek

